# Bacterial genotypic and patient risk factors for adverse outcomes in *Escherichia coli* bloodstream infections: a prospective molecular-epidemiological study

**DOI:** 10.1101/2021.06.18.21258557

**Authors:** Elita Jauneikaite, Kate Honeyford, Oliver Blandy, Mia Mosavie, Max Pearson, Farzan A. Ramzan, Matthew J. Ellington, Julian Parkhill, Céire E Costelloe, Neil Woodford, Shiranee Sriskandan

**Author notes:** Corresponding author: Prof Shiranee Sriskandan). These authors contributed equally (Elita Jauneikaite, Kate Honeyford).

## Abstract

**Background:** *Escherichia coli* bloodstream infections have increased rapidly in the UK, for reasons that are unclear. The relevance of highly fit, or multi-drug resistant lineages such as ST131 to overall *E. coli* disease burden remains to be fully determined. We set out to characterise the prevalence of *E. coli* multi-locus sequence types (MLST) and determine if these were associated with adverse outcomes in an urban population of *E. coli* bacteraemia patients.

**Methods:** We undertook whole genome sequencing of *E. coli* blood isolates from all patients with diagnosed *E. coli* bacteraemia in north-west London from July 2015 to August 2016 and assigned multi-locus sequence types to all isolates. Isolate sequence types were linked to routinely collected antimicrobial susceptibility, patient demographic, and clinical outcome data to explore relationships between the *E. coli* sequence types, patient factors, and outcomes.

**Findings:** A total of 551 *E. coli* genomes were available for analysis. More than half of these cases were caused by four *E. coli* sequence types: ST131 (21%), ST73 (15%), ST69 (9%) and ST95 (8%). *E. coli* genotype ST131-C2 was associated with non-susceptibility to quinolones and third-generation cephalosporins, and also to amoxicillin, augmentin, gentamicin and trimethoprim. An association between the ST131-C2 lineage and longer length-of-stay was detected, although multivariable regression modelling did not demonstrate an association between *E. coli* sequence type and mortality. However, a number of unexpected associations were identified, including gentamicin non-susceptibility, ethnicity, and sex that might influence mortality and length-of-stay, requiring further research.

**Interpretation:** Although *E. coli* sequence type was associated with antimicrobial non-susceptibility patterns and length-of-stay, we did not find that *E. coli* sequence type was associated with increased mortality. Where ST131 is prevalent, caution is required when pairing beta-lactam agents with gentamicin or using single agent aminoglycosides.

**Funding:** UK NIHR HPRU, Wellcome Trust, Rosetrees Trust, Stoneygate Trust.

**Research in context:** *Evidence before this study:* *E. coli* is the leading cause of bacteraemia in adults, with consequent major impact on patient morbidity and mortality, as well as cost of care. Identification of microbial and patient factors that contribute to severity from *E. coli* bacteraemia could inform clinical guidelines and improve outcomes. We searched PubMed for articles published in English between 1^st^ of Jan 1999 to 3^rd^ of May 2021 using the following terms: (“Escherichia coli” OR “E. coli”) AND (“bacteraemia” OR “bloodstream infection”) AND (“genotype” OR “multi-locus sequence typing” OR “MLST”). We screened titles, abstracts, and bibliographies of relevant articles to identify reports that examine microbial factors that influence outcome. Previous studies have examined the association between a narrow range of *E. coli* lineages carrying specific antimicrobial resistance genes and prior antibiotic consumption, patient comorbidities, and mortality. Only two previous studies have examined the association of a wider range of *E. coli* sequence types (via MLST or whole genome sequencing) with clinical disease phenotype. We provide an integrated observational analysis of *E. coli* sequence types, patient demographic data, and disease outcomes.

*Added value of this study:* We provide whole genome sequences from an un-biased collection of 551 *E. coli* strains causing consecutive bloodstream infections (BSI) in a one-year period in London. The study confirms ST131 to be the single leading BSI-causing genotype, albeit accounting for just 21% of infections. Only half of these were from the so-called multi-drug resistant ST131-C2 lineage however, suggesting factors other than antimicrobial resistance alone contribute to fitness of ST131, and other currently dominant lineages. The study provides a valuable resource to link genome sequence to antimicrobial resistance phenotypes and evaluates the contribution of both bacterial sequence type and patient factors to disease outcome. *E. coli* ST131 subclade C2 was associated with non-susceptibility to multiple antibiotics as well as a longer length-of-stay, underlining a value in sequence-type surveillance, but we did not detect an association with mortality. Multi-variate analysis unexpectedly identified ethnicity as a potential contributor to 90-day mortality, mandating further investigation, while microbial gentamicin resistance was also linked to adverse outcome.

*Implications of all available evidence:* *E. coli* bloodstream infections continue to rise in the UK, despite implemented strategies aimed at reducing *E. coli* invasive infections and antimicrobial resistance. Certain *E. coli* lineages are more likely to be multidrug-resistant or associated with specific infection types, however a more detailed understanding of changes in these pathogenic lineages over time in vulnerable populations is needed. Although multi-drug resistant lineages such as ST131-C2 contribute to prolonged length-of-stay, factors other than *E. coli* genotype may have a greater role to play in final outcome. While co-morbidities play a major role in mortality from *E. coli* bacteraemia, other demographic factors require attention. A more comprehensive analysis of associations between *E. coli* genotype, patient factors, antimicrobial resistance phenotypes and clinical outcomes could inform prescribing guidelines for both urinary tract and invasive *E. coli* infections. Such information will further serve as a potential driver for developing and implementing lineage-specific *E. coli* vaccines in the future.

## Introduction

*Escherichia coli* is, by far, the most common causative organism in bloodstream infections (BSIs) [1] and its incidence is increasing. Between 2014-18 a 27·2% increase in *E. coli* BSIs was recorded in England, Wales and Northern Ireland, rising from 55·2/100,000 to 70·7/100,000 cases [2]. In-hospital mortality of *E. coli* BSI is reported to be 13-25% [3–5] and prolonged length-of-stay is common [5–7]. Thus, with over 40,000 cases per year [8], the overall burden of *E. coli* BSIs on individuals and healthcare is considerable. Additionally, non-susceptibility to several commonly-used antimicrobial agents has increased among both community- and hospital-onset *E. coli* blood isolates [7,9].

While many studies have described the factors that may influence outcome from sepsis and *E. coli* BSIs [6,7], few have examined whether strain sequence type is relevant. Extraintestinal pathogenic *E. coli* (ExPEC) form a subgroup of *E. coli* that have sufficient virulence to cause urinary and bloodstream infections [10]. The most common disease-causing clones of ExPEC in the UK have been reported to be ST131, ST73, ST95, ST69 and ST12 [11]. Globally, interest has focused on emergence of the ST131 *E. coli* lineage, in particular the multidrug-resistant clade ST131-C2/H30-Rx, which is associated with production of extended-spectrum beta-lactamases (ESBL) and resistance to fluoroquinolones [12–14]. Although ST131 to date has not outcompeted other common clones [15], the emergence of the *E. coli* ST131-*H30*-Rx (also known as ST131-C2) sub-lineage has reportedly been associated with more severe infections [16] and characterised as highly pathogenic [14]. We conducted a one-year prospective study to investigate the burden of infection due to specific *E. coli* genotypes and to determine if *E. coli* genetic background was associated with adverse outcome in bacteraemia patients.

## Materials and Methods

### Case definitions, demographic and clinical outcome data retrieval

Between July 2015 and August 2016, we prospectively collected *E. coli* isolates causing bloodstream infections submitted to the diagnostic laboratory of a large NHS teaching Trust in North West London, serving a population of ∼2,000,000 people including adjacent NHS Trust hospitals.

*E. coli* bacteraemia cases were identified when a positive blood culture yielded growth of *E. coli*. Any *E. coli* bacteraemia events recorded within 30 days of an earlier *E. coli*-positive blood culture were considered duplicates and not included in the analysis; those recorded >30 days after were considered new infections and were included in the study as cases. *E. coli* bacteraemia cases were considered to be hospital-onset (HO) if the *E. coli*-positive blood culture was taken after the second calendar day of admission to hospital; or community-onset (CO) if the *E. coli*-positive blood culture was taken on the first or second day of admission [17]. The following demographic information was collected using an electronic patient administration system (PAS): age, sex, ethnicity, co-morbidities, post-infection length-of-stay (Figure S1). A modified Elixhauser comorbidity score was calculated for each case [18]. The primary outcome was all-cause mortality within 7, 30 and 90 days of positive blood culture date, derived by linking cases to Office for National Statistics (ONS) data. Post-infection length-of-stay was determined from hospital PAS data but was highly skewed. A binary indicator of prolonged length-of-stay (≥7; https://improvement.nhs.uk/resources/reviewing-str) was used as a secondary patient outcome. Detailed demographic and length-of-stay data were not available for patients admitted to the adjacent NHS Trust hospitals.

### Antibiotic susceptibility testing and multi-locus sequence typing

Antibiotic susceptibility testing results for *E. coli* blood isolates were obtained from the Microbiology Data warehouse. Testing was carried out as part of the routine clinical care following European Committee on Antimicrobial Susceptibility Testing (EUCAST) methodology (https://eucast.org/ast_of_bacteria/) for amoxicillin, augmentin, trimethoprim, fluoroquinolones (ciprofloxacin), third-generation cephalosporins (ceftazidime, cefotaxime and ceftriaxone), carbapenems (imipenem, meropenem, ertapenem) and aminoglycosides (gentamicin). For analysis, intermediate and resistant isolates were combined and classified as ‘non-susceptible’. Isolates which were non-susceptible to three classes of antimicrobial agents (ciprofloxacin, third-generation cephalosporins and gentamicin) were classed as multidrug-resistant (MDR). Amoxicillin, augmentin, and trimethoprim susceptibility data were not available for patients admitted to adjacent NHS Trust hospitals. *E. coli* isolates were processed for whole genome sequencing, and genotypes based on multi-locus sequence types (MLST) were determined as described in Supplementary Methods.

### Statistical analyses

Chi-squared and Fisher’s exact tests were used to assess associations between *E. coli* sequence type, antibiotic susceptibility results and patient demographic data (sex and age). Multiple logistic regression was used to model the outcomes, mortality within 7, 30 and 90 days and length-of-stay. As mortality is a rare outcome, the Firth method [19,20] was used in mortality models to reduce the small sample bias in Maximum Likelihood Estimation. *E. coli* genotype was included as a categorical model in all models. All patient characteristics were considered as potential confounders for both outcomes and were included in all models. To isolate potential confounding introduced by antibiotic susceptibility in *E. coli* sequence types, we included antibiotic susceptibility results for each antibiotic one at a time in models. All the tests were two-sided, and p<0.05 was considered statistically significant. Data processing and analyses were performed using R studio (http://www.r-project.org).

### Ethics statement

The collection of microbial isolates and linkage to routinely obtained healthcare data prior to anonymisation was approved by an NHS Research Ethics Committee (West London Research Ethics Committee, formerly Hammersmith and Queen Charlotte’s & Chelsea Research Ethics Committee). Ethics approval reference number 06/Q0406/20.

## Results

### Description of one-year E. coli bloodstream infections in north-west London

A total of 551 *E. coli* bacteraemia cases, where *E. coli* isolates were available for analysis, were detected between July 2015 and August 2016 (Figure S1). Basic demographic information (age and sex) was available for 537 patients (Figure S1). Of these, 43% were male (n=233) and 57% were female (n=304). Sixty percent of *E. coli* bacteraemia cases were ≥65 years old (n=321), with more female patients represented in this age group than male patients (Table S1). Genomic MLST analysis identified 114 different MLST among *E. coli* blood isolates, of which 21% belonged to ST131 (n=115), 15% to ST73 (n=77), 9% to ST69 (n=51) and 8% to ST95 (n=45). As such, over half of all isolates belonged to ST131, ST73, ST69 or ST95. Although ST131 was the single largest lineage among bacteraemia isolates, within the ST131 lineage there were 60/115 isolates that belonged to the ST131-C2 sub-lineage (11·2% of total) and the remaining 55 (10·2% of the total) isolates from “other” ST131 sub-lineages (including A, B and C1).

### E. coli genotypes (MLST) and antimicrobial susceptibility

Results of routinely conducted antibiotic susceptibility testing to five key antibiotic groups (fluoroquinolones, third-generation cephalosporins, carbapenems, gentamicin and tazocin) were available for 537 isolates, while susceptibilities to amoxicillin, augmentin and trimethoprim were available for 249 isolates (Figure S1). From the five antibiotic groups tested, 26·7% of isolates (n=143/537) were non-susceptible to fluoroquinolones, 19·7% of isolates (n=106/537) were non-susceptible to third-generation cephalosporins, and 15·1% (n=81/537) were non-susceptible to gentamicin (Table S2). Non-susceptibility to amoxicillin (63·9%, n=159/249), trimethoprim (44·6%, n=111/249) and augmentin (30·5%, n=76/249) was frequently noted. Non-susceptibility to any of the antibiotics tested (except for carbapenems due to low isolate numbers tested) was significantly associated with the *E. coli* MLST (p<0.001), while only non-susceptibility to amoxicillin was not associated with the MLST (p>0.05) (Table S2). As expected, ST131-C2 showed the highest levels of non-susceptibility to all antibiotics tested (Table S2). ST131-C2 accounted for approximately one third of isolates that were non-susceptible to third-generation cephalosporins and quinolones; and accounted for approximately one quarter of isolates that were non-susceptible to gentamicin, tazocin, or augmentin. Although non-susceptibility to trimethoprim was observed frequently, in 44·6% of all *E. coli* isolates tested (111/249), it was notable that 81·5% of ST131-C2 isolates (n=22/27) were non-susceptible to trimethoprim (Table S2).

### Association between E. coli MLST and case characteristics

*E. coli* MLST was linked to clinical demographic data in 300 cases. Initial analysis indicated that ST69 and ST95 isolates were more frequently identified in younger (<65 yrs) female patients, while ST131 isolates were more likely to affect male patients (Table 1). There was, however, no clear association between specific *E. coli* MLST and age or ethnicity.

**Table 1.** Summary of patient characteristics and association with specific *E. coli* sequence type (n=300 patients).

|  | ST131-C2<br>(n=36) | ST131-other<br>(n=28) | ST69<br>(n=31) | ST73<br>(n=43) | ST95<br>(n=21) | Other STs<br>(n=114) | Total<br>(n=300) |
| --- | --- | --- | --- | --- | --- | --- | --- |
| **Sex** |  |  |  |  |  |  |  |
| **Male** | 20 (55.6%) | 16 (57.1%) | 8 (25.8%) | 20 (46.5%) | 8 (38.1%) | 65 (48.6%) | 144 (48.0%) |
| **Female** | 16 (44.4%) | 12 (42.9%) | 23 (74.2%) | 23 (53.5%) | 13 (61.9%) | 69 (51.4%) | 156 (52.0%) |
| **Age** |  |  |  |  |  |  |  |
| **< 65** | 11 (30.6%) | 14 (50.0%) | 17 (54.8%) | 19 (44.2%) | 9 (42.9%) | 61 (43.3%) | 131 (43.7%) |
| **65-74** | 12 (33.3%) | 6 (21.4%) | 4 (12.9%) | 7 (16.3%) | 4 (19.0%) | 35 (24.8%) | 68 (22.7%) |
| **75-84** | 6 (16.7%) | 6 (21.4%) | 8 (25.8%) | 8 (18.6%) | 6 (28.6%) | 33 (23.4%) | 67 (22.3%) |
| **85+** | 7 (19.4%) | 2 (7.1%) | 2 (6.5%) | 9 (20.9%) | 2 (9.5%) | 12 (8.5%) | 34 (11.3%) |
| **Ethnicity** |  |  |  |  |  |  |  |
| **White** | 16 (44.4%) | 10 (35.7%) | 16 (51.6%) | 28 (65.1%) | 10 (47.6%) | 65 (46.1%) | 145 (48.3%) |
| **Asian** | 9 (25.0%) | 5 (17.9%) | 4 (12.9%) | 2 (4.7%) | 2 (9.5%) | 19 (13.5%) | 41 (16.7%) |
| **Black** | 4 (11.1%) | 5 (17.9%) | 3 (9.7%) | 2 (4.7%) | 2 (9.5%) | 17 (12.1%) | 33 (11.0%) |
| **Any Other** | 6 (16.7%) | 2 (7.1%) | 5 (16.1%) | 4 (9.3%) | 3 (14.3%) | 24 (17.0%) | 44 (14.7%) |
| **Not Given** | 1 (2.8%) | 6 (21.4%) | 3 (9.7%) | 7 (16.3%) | 4 (19.0%) | 16 (11.3%) | 37 (12.3%) |
| **Elixhauser score** |  |  |  |  |  |  |  |
| **<0** | 0 (0%) | 0 (0%) | 0 (0%) | 3 (7.0%) | 0 (0%) | 7 (5.0%) | 10 (3.3%) |
| **Zero** | 11 (30.6%) | 3 (10.7%) | 11 (35.5%) | 8 (18.6%) | 6 (28.6%) | 30 (21.3%) | 69 (23.0%) |
| **1 to 5** | 8 (22.2%) | 6 (21.4%) | 9 (29.0%) | 8 (18.6%) | 2 (9.5%) | 29 (20.6%) | 62 (20.7%) |
| **6 to 13** | 9 (25.0%) | 11 (39.3%) | 6 (19.4%) | 18 (41.9%) | 10 (47.6%) | 36 (25.5%) | 90 (30.0%) |
| **≥14** | 8 (22.2%) | 8 (28.6%) | 5 (16.1%) | 6 (14.0%) | 3 (14.3%) | 39 (27.7%) | 69 (23.0%) |
| **Onset of infection** |  |  |  |  |  |  |  |
| **Hospital** | 15 (41.7%) | 11 (39.3%) | 7 (22.6%) | 8 (18.6%) | 2 (9.5%) | 32 (22.7%) | 75 (25.0%) |
| **Community** | 20 (55.6%) | 17 (60.7%) | 21 (67.7%) | 35 (81.4%) | 19 (90.5%) | 104 (73.8%) | 225 (75.0%) |

Elixhauser co-morbidity scores exceeded 5 for 53·6% of the patients (Table 1). We observed that patients infected with ST131, ST73 and ST95 were more likely to have an Elixhauser score >6, compared to patients infected with other STs (Table 1). Within the ST131 lineage, patients infected with “ST131-other” isolates were more likely to have higher Elixhauser score than those infected with ST131-C2 isolates (Table 1).

Based on onset of *E. coli* bacteraemia in relation to time of admission to hospital, 75% of patients had community-onset *E. coli* bacteraemia (Table 1). *E. coli* MLST were, in general, proportionately distributed among community- and hospital-onset cases, except for ST131 isolates that were disproportionately associated with hospital-onset cases; 40% of ST131 isolates were hospital-onset (Table 1). Overall, the ST131 lineage accounted for 34·6% of all hospital-onset cases compared with 16·4% of all community-onset cases. In contrast, a majority (90%) of bacteraemia cases caused by ST95 were found to be community-onset (Table 1).

### Association between E. coli MLST and patient outcome

Over one-third (34%) of study patients with *E. coli* bacteraemia died within one-year: 7% died within 7 days of infection, 11% died within 30 days and 16% within 90 days (Table 2). There was no association between *E. coli* MLST and death within 7 days or 30 days in the cohort tested, however, analysis may have been affected by low numbers within each *E. coli* genotype group, and mortality being lower than predicted, precluding further analysis at the early time points (Table 2). Logistic regression analysis showed increased risk of death within 90 days for patients infected with *E. coli* genotype ‘ST131-other’ when compared to all other STs (OR:2.58; 95%CI:1.04, 6.19). However, once the model was adjusted for patient characteristics, no evidence for any association between *E. coli* genotype and 90-day mortality was found (Table 3). In our study, age and sex did not influence risk of 90-day mortality, although comorbidity, as described by an Elixhauser score of >14, was highly influential. Unexpectedly, we found that *E. coli* non-susceptibility to gentamicin was associated with increased odds of death within 90 days (OR:3·32, 95%CI:1·17, 9·72) after adjusting for patient characteristics and *E. coli* MLST (Table 3). Mortality at all time points was higher in patients infected by isolates that were non-susceptible to gentamicin (7 days, 12·8%; 30 days, 15·4%; 90 days, 25·6%), compared with patients with isolates that were susceptible, (7 days, 6·1%; 30 days, 10·3%; 90 days, 14·2%) although numbers at the earlier time points were too low for inclusion in comparative analysis. Further analysis also suggested that patients of black ethnicity had increased odds of mortality within 90 days compared to patients of white ethnicity: Those infected with *E. coli* strains that were non-susceptible to gentamicin or tazocin were associated with increased odds of death within 90 days (OR: 2·98, 95%CI:1·03,8·56 and OR:3·09, 95%CI:1·02,9·26, respectively; Table 3).

**Table 2.** Summary of results for testing association between *E. coli* sequence type and patient outcome (n=300 patients).

| Outcome | ST131-C2<br>(n=36) | ST131-other<br>(n=28) | ST69<br>(n=31) | ST73<br>(n=43) | ST95<br>(n=21) | Other STs<br>(n=141) | Total<br>(n=300) |
| --- | --- | --- | --- | --- | --- | --- | --- |
| **Mortality** |  |  |  |  |  |  |  |
| **Within 7 days n (%)** | 1<br>(2.8%) | 4<br>(14.3%) | 1<br>(3.2%) | 3<br>(7.0%) | 1<br>(4.8%) | 11<br>(7.8%) | 21<br>(7.0%) |
| **Within 30 days n (%)** | 2<br>(5.6%) | 6<br>(21.4%) | 2<br>(6.5%) | 3<br>(7.0%) | 2<br>(9.5%) | 18<br>(12.8%) | 33<br>(11.0%) |
| **Within 90 days n (%)** | 4<br>(11.1%) | 10<br>(35.7%) | 3<br>(9.7%) | 3<br>(7.0%) | 2<br>(9.5%) | 25<br>(17.7%) | 47<br>(15.7%) |
| **Length-of-stay post infection** |  |  |  |  |  |  |  |
| **Median (IQR)** | 18.5<br>(11.25, 27.0) | 12.5<br>(6, 28.75) | 9<br>(7, 15) | 7<br>(5, 13) | 11<br>(6, 20) | 9<br>(11.25, 27) | 10<br>(6, 20) |
| **(min, max)** | (1, 75) | (1, 50) | (1, 62) | (0, 90) | (1, 64) | (0, 91) | (1, 62) |
| **Patients who have stayed 7 or more days** | 32<br>(94.1%) | 20<br>(71.4%) | 22<br>(78.6%) | 25<br>(58.1%) | 14<br>(66.7%) | 91<br>(66.9%) | 204<br>(68%) |

**Table 3.**
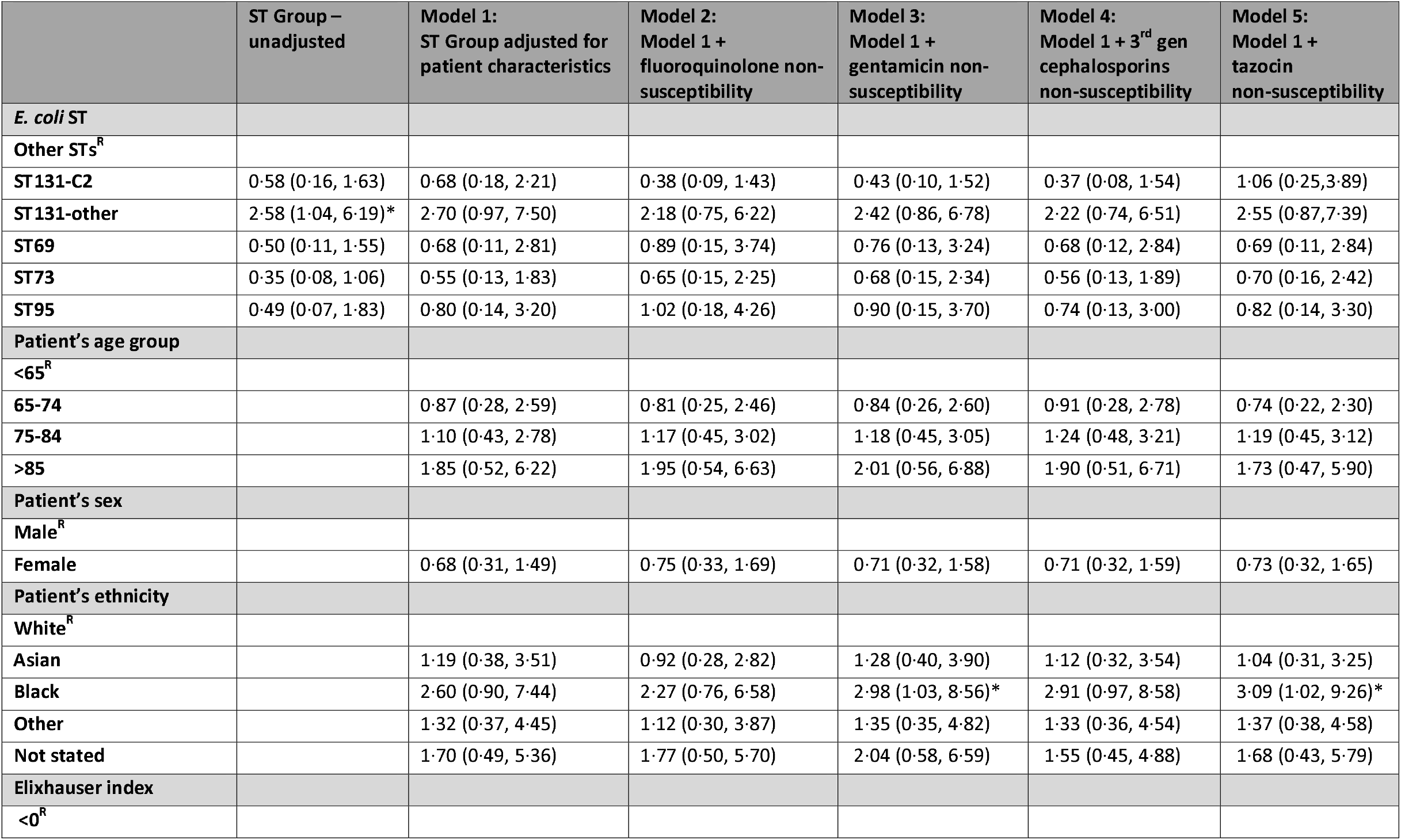
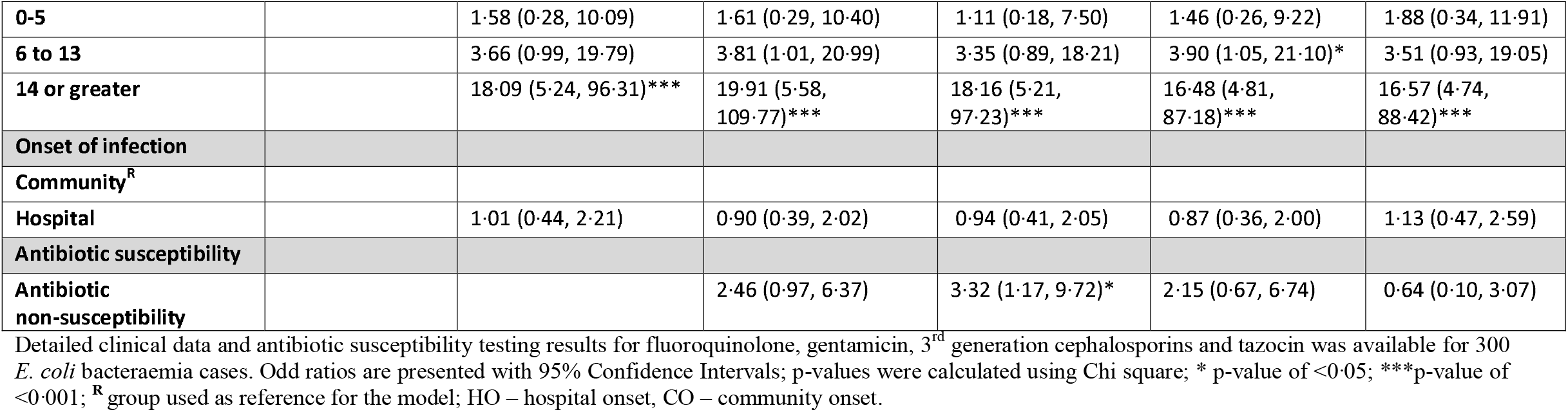
Multiple regression modelling of association between *E. coli* genotype, patient characteristics, antibiotic resistance and mortality within 90 days.

Median post infection length-of-stay (PILOS) was found to be 10 days for *E. coli* bacteraemia patients (Table 2). Overall patients with bacteraemia caused by ST131 (irrespective of clade) were more likely to stay longer in hospital, with longest PILOS for patients infected with ST131-C2 isolates (median 18.5 days) and ST131-other isolates (median 12.5 days) (Table 2). Unadjusted logistic regression suggested that patients infected with ST131-C2 had six times greater odds of having an extended length-of-stay (OR:6·46 95%CI: 2·02 to 32·64) and patients infected with ST69 had a three-fold increase in odds of a long length-of-stay (OR:2·85 95%CI:1·07 to 8·52) compared with patients who were infected with ‘other’ ST *E. coli* (Table 4). We did not find evidence of a specific association between antibiotic non-susceptibility and length-of-stay. Female patients had a three-fold lower risk of an extended length-of-stay compared with male patients (OR:0·30 95%CI:0·16, 0·55) (Table 4). Patients with hospital-onset infection had higher odds of an extended length-of-stay (OR:2·69 95%CI:1·28, 6·08) although these patients may have required treatment for the condition that initiated the original admission to hospital (Table 4). When we examined the smaller subset of patients for whom additional antimicrobial susceptibility data were available, we were unable to detect any association between non-susceptibility to trimethoprim, amoxicillin or augmentin and either PILOS or death within 90 days (Tables S3-S4).

**Table 4.** Multiple regression modelling of association between *E. coli* genotype, patient characteristics, antibiotic resistance and length-of-stay (>7 days). Detailed clinical data and antibiotic susceptibility testing results for fluoroquinolone, aminoglycoside, 3^rd^ generation cephalosporins and tazocin from Local NHS Trust was available for 300 *E. coli* bacteraemia cases.

|  | ST Group –<br>unadjusted | Model 1:<br>ST Group adjusted<br>for patient<br>characteristics | Model 2:<br>Model 1 +<br>fluoroquinolone<br>non-susceptibility | Model 3:<br>Model 1 +<br>aminoglycoside<br>non-susceptibility | Model 4:<br>Model 1 + 3<sup>rd</sup> gen<br>cephalosporins<br>non-susceptibility | Model 5:<br>Model 1 +<br>tazocin<br>non-susceptibility |
| --- | --- | --- | --- | --- | --- | --- |
| *E. coli* genotype |  |  |  |  |  |  |
| Other STs<sup>R</sup> |  |  |  |  |  |  |
| ST131-C2 | 6.46 (2.02, 32.64)*** | 6.06 (1.75, 32.15)** | 4.81 (1.23, 27.10)* | 5.80 (1.63, 31.13)** | 8.93 (1.85, 89.15)** | 4.54(1.25, 24.94)* |
| ST131-other | 1.20 (0.51, 3.01) | 0.81 (0.32, 2.17) | 0.71 (0.27, 1.95) | 0.77 (0.30, 2.11) | 0.79 (0.30, 2.17) | 0.84(0.32, 2.39) |
| ST69 | 1.72 (0.70, 4.76) | 2.85 (1.07, 8.52)* | 3.06 (1.14, 9.20)* | 2.97 (1.10, 8.95)* | 3.13 (1.17, 9.37)* | 2.82(1.04, 8.59)* |
| ST73 | 0.69 (0.34, 1.39) | 0.70 (0.33, 1.53) | 0.79 (0.36, 1.77) | 0.84 (0.38, 1.86) | 0.79 (0.35, 1.77) | 0.82(0.35, 1.96) |
| ST95 | 0.96 (0.38, 2.61) | 1.29 (0.46, 3.85) | 1.38 (0.49, 4.18) | 1.31 (0.46, 3.96) | 1.40 (0.50, 4.22) | 1.15(0.40, 3.54) |
| Patient's age<br>group |  |  |  |  |  |  |
| <65<sup>R</sup> |  |  |  |  |  |  |
| 65-74 |  | 1.43 (0.69, 3.04) | 1.46 (0.70, 3.12) | 1.41 (0.68, 3.00) | 1.40 (0.66, 3.02) | 1.48(0.70, 3.23) |
| 75-84 |  | 1.61 (0.74, 3.57) | 1.67 (0.77, 3.71) | 1.75 (0.80, 4.00) | 1.58 (0.72, 3.55) | 1.87(0.82, 4.46) |
| >85 |  | 1.64 (0.65, 4.38) | 1.87 (0.72, 5.16) | 1.79 (0.69, 4.95) | 1.53 (0.58, 4.29) | 1.84(0.68, 5.33) |
| Patient's sex |  |  |  |  |  |  |
| Male<sup>R</sup> |  |  |  |  |  |  |
| Female |  | 0.30 (0.16, 0.55)*** | 0.33 (0.18, 0.62)*** | 0.29 (0.15, 0.54)*** | 0.29 (0.15, 0.53)*** | 0.30(0.15, 0.57)*** |
| Patient's ethnicity |  |  |  |  |  |  |
| White<sup>R</sup> |  |  |  |  |  |  |
| Asian |  | 1.26 (0.52, 3.26) | 1.15 (0.47, 3.02) | 1.56 (0.62, 4.24) | 1.14 (0.46, 3.01) | 1.53(0.58, 4.36) |
| Black |  | 1.20 (0.45, 3.46) | 1.16 (0.43, 3.35) | 1.24 (0.46, 3.61) | 1.03 (0.37, 3.05) | 0.93(0.33, 2.80) |
| Other |  | 0.78 (0.34, 1.83) | 0.77 (0.34, 1.81) | 0.78 (0.34, 1.84) | 0.68 (0.28, 1.64) | 0.81(0.34, 1.98) |
| **Not stated** |  | 0.78 (0.33, 1.87) | 0.75 (0.32, 1.80) | 0.79 (0.34, 1.90) | 0.79 (0.34, 1.90) | 0.74(0.30, 1.85) |
| **Elixhauser index** |  |  |  |  |  |  |
| **< 0<sup>R</sup>** |  |  |  |  |  |  |
| **0-5** |  | 2.15 (0.98, 4.79) | 2.10 (0.96, 4.68) | 2.09 (0.95, 4.70) | 1.93 (0.87, 4.37) | 2.08 (0.93, 4.79) |
| **6 to 13** |  | 2.71 (1.28, 5.87)** | 2.70 (1.27, 5.91)** | 2.91 (1.36, 6.41)** | 2.43 (1.11, 5.44)* | 3.36 (1.52, 7.66)** |
| **14 or greater** |  | 2.02 (0.87, 4.80) | 2.06 (0.89, 4.89) | 1.91 (0.82, 4.56) | 1.85 (0.79, 4.46) | 2.27 (0.94, 5.67) |
| **Onset of infection** |  |  |  |  |  |  |
| **Community onset<sup>R</sup>** |  |  |  |  |  |  |
| **Hospital onset** |  | 2.69 (1.28, 6.08)** | 2.51 (1.18, 5.71)* | 2.62 (1.24, 5.92)* | 2.55 (1.17, 5.95)* | 2.55 (1.15, 6.10)* |
| **Antibiotic susceptibility** |  |  |  |  |  |  |
| **Antibiotic non-susceptibility** |  |  | 1.42 (0.65, 3.20) | 1.14 (0.41, 3.44) | 1.12 (0.41, 3.18) | 0.53 (0.13, 2.31) |

Odd ratios are presented with 95% Confidence Intervals; p-values were calculated using Chi square; \* p-value of <0.05; \*\*\*p-value of <0.001; <sup>R</sup> group used as reference for the model; HO – hospital onset, CO – community onset.

## Discussion

Our study is one of the largest to comprehensively examine the association between *E. coli* genotype and bloodstream infection, examining over 500 consecutive unselected cases of bacteraemia. Over one fifth of cases could be attributed to isolates of the ST131 lineage, around half of which belonged to the multidrug-resistant sub-clone ST131-C2 (also known as ST131-*H30Rx*). It has been reported that successful spread of ST131 clades was due to gain of virulence-associated genes, followed by the acquisition of specific antibiotic resistance [21]. Although the ST131-C2 clade is reported to be highly pathogenic [14] we did not find clinical-epidemiological evidence to support this in our patient population when mortality was used as a surrogate for virulence and pathogenicity; the most influential factor was patient comorbidity.

In contrast to mortality, when extended length of hospital stay was considered, certain *E. coli* genotypes, including ST131-C2, were strongly associated with prolonged length-of-stay, after adjusting for patient characteristics. This observation could potentially be explained by the non-susceptibility of ST131-C2 isolates to available oral antimicrobials that would otherwise be used to expedite patient discharge after initial intravenous therapy. Interestingly, bloodstream infections due to ST69 were also associated with a prolonged length-of-stay. Based on the data available, there was no evidence that non-susceptibility to multiple antibiotics alone was associated with extended length-of-stay although our ability to detect such a difference may have been limited by study size.

A number of other studies have examined the factors that influence outcome following *E. coli* bacteraemia. Most have identified the importance of comorbidity and age on mortality. Some have identified factors that are amendable to intervention such as timing of effective antibiotics [22,23]. Only two have examined the role of bacterial sequence type, but failed to identify an association with adverse outcome such as mortality [16,24]. The role of strain genotype in mortality in invasive bacterial infections caused by other species is seen to reflect a salient role for the bacterium in pathogenesis[25]. The lack of identifiable link to any one genotype in the current study may reflect that *E. coli* is largely an opportunistic pathogen, and emergent lineages may simply represent strain types that are well-adapted to colonise the gut or cause disease in the elderly and those with comorbidity. *E. coli* is the leading cause of bloodstream infection in the UK and will account for a majority of cases that are designated as sepsis; factors that influence sepsis outcome will therefore be dominated by the largely host-related factors that impact *E. coli* outcome[6,7].

Genotyping of the *E. coli* isolates in this study also provided insight into the MLST-specific associations with markers of antimicrobial resistance. A retrospective study undertaken in north-west London highlighted an unexplained increase in quinolone non-susceptibility in *E. coli* bacteraemia isolates 2011-2015 [7]. The ST131-C2 sub-lineage is known to be associated with non-susceptibility to quinolones, third-generation cephalosporins and gentamicin [26] and this was confirmed in our study, as was non-susceptibility to amoxicillin (85%), and augmentin (67%). These antibiotics are preferentially used when treating *E. coli* bloodstream infections, however the emergence of ST131-C2 and other ESBL-producing Gram-negative bacteria has led to increasing empiric use of carbapenem agents or combination regimens that include aminoglycosides. Our frequent finding of gentamicin non-susceptibility in ST131 isolates (34%) points to a risk in the latter approach in populations where ST131-C2 is present. Although we did not specifically examine amikacin susceptibility in this study, it would appear that alternate regimens that include amikacin would be safer [27]. Our wider phenotypic testing against antimicrobials that are used in the community, for urinary tract rather than blood infections, further demonstrated that a striking proportion (>80%) of ST131-C2 isolates are trimethoprim resistant. Trimethoprim was recommended for empiric management of uncomplicated urinary tract infection until 2018 and we speculate that this may have contributed to population emergence of ST131 in those with frequent urosepsis and, in part, contributed to the predominance of ST131-C2 in bacteraemia cases.

Multivariable regression analysis unexpectedly highlighted an association between gentamicin resistance and increased odds of 90-day mortality (OR:3·32, 95% CI:1·17 to 9·72). Mortality at earlier time points also appeared greater in this group of patients, although the low numbers preclude statistical evaluation (at 7 days, gentamicin susceptible 6·1% versus non-susceptible 12·8%; at 30 days, gentamicin susceptible 10·3% versus non-susceptible 15·4%). Aminoglycosides are frequently used as adjuncts to beta-lactam antimicrobials in empiric management of suspected sepsis, to avoid use of broader spectrum agents such as carbapenems. In our study, 27% of isolates resistant to augmentin were also resistant to gentamicin, and 47% isolates resistant to cephalosporins were resistant. Empiric single agent aminoglycoside therapy has previously been proposed as treatment for suspected urinary tract infection, although there is no evidence to support this approach in the setting of sepsis [28]. In either case, the proposed regimes will fail to achieve microbiological clearance of an aminoglycoside resistant ESBL-producing *E. coli* infection. Our observation of a three-fold increased risk of death in such cases underlines the importance of local surveillance of resistance patterns and adjustment of protocols accordingly, for example use of amikacin rather than gentamicin in a dual-agent regimen, and ensuring antibiotic regimens are initiated and adjusted promptly[22].

Unexpectedly, the multivariable models also identified black ethnicity as a predictor of mortality; this could not be explained by patient comorbidities or specific *E. coli* lineages. It is feasible that ethnicity may play a role in late recognition of bacteraemia with consequent impact on management and therefore sepsis outcomes. Ethnicity is a complex trait that has frequently been associated with increased sepsis mortality in studies undertaken in North America [29]. In our study, black ethnicity was associated with a mortality of 6% at 7d, 18·2% at 30 days, and 33·3% at 90 days, in contrast to white ethnicity where mortality was 6·9%. 8·3% and 11·7% at the same time points respectively albeit that numbers were too low at earlier time points to analyse. We do not know if similar data exist for other infections, or other types of hospital admission in the UK, however the recent COVID-19 pandemic has highlighted the role of inequality in healthcare outcomes [30]. Socio-economic status and travel history were not collected in our study, and we cannot rule out the possibility that specific groups may be over-represented among patients from long term care facilities or specific healthcare settings, for example dialysis or haematology. Our findings mandate more detailed study of the impact that ethnicity and other social factors might play in *E. coli* bacteraemia outcome within the UK.

The analysis also revealed that female patients were three times less likely to have a prolonged length-of-stay compared to male patients, even when adjusted for confounding factors, resonating with findings in an earlier national retrospective cohort study [5]. Length-of-stay was also markedly prolonged in those patients with hospital-onset *E. coli* bloodstream infections, as found in a previous retrospective study carried out in our NHS Trust [7]: The models applied in the current work accounted for hospital onset and comorbidities, but not the original reason for hospital admission which may in itself be the main driver for prolonged length-of-stay. There may also be important associations between reason for admission, onset of infection and gender. Larger samples and more detailed information would allow these more complex associations and possible interactions to be examined.

There are limitations to our study. The study took place over one-year and was based in an urban and socially diverse area in London, hence, our findings might not be relevant to other settings. The study was under-powered to detect significant differences in mortality between *E. coli* genotype as the observed mortality at 30d was much lower than predicted; nonetheless, it was clear that ST131-clade C alone was not hypervirulent. Ethnicity is self-reported with missing data in many cases, and we had no information on the social status of the patients to adjust for in the analysis of association between *E. coli* genotype, severity of the infection and ethnicity. Information on other risk factors such as previous antibiotic exposure or transfer from other facilities (such as elderly care or long-term care facilities) was not available to include in our analyses. Finally, although our study included over 500 cases of bloodstream infection and isolates, we only had full outcome data for 300 patients, limited to the local NHS Trust and antimicrobial susceptibility testing was limited to the phenotypic tests reported by the local diagnostic laboratory.

## Conclusions

One in five *E. coli* bloodstream infections occurring in north-west London arise from the *E. coli* ST131 lineage, half of which can be assigned to multi-drug resistant clade ST131-C2. There was no evidence that clade ST131-C2 was linked to increased mortality, indeed, there was a trend for other ST131 infections to have higher mortality. The main determinants of bacteraemia outcome were comorbidity-related, while bacterial MLST was strongly linked to antimicrobial resistance phenotype and length-of-stay. Future work should aim to recruit a larger cohort of bloodstream infection cases and isolates to examine the potential roles of ethnicity, strain type and resistance phenotype with outcome.

## Data Availability

Whole genome sequencing reads were deposited in European Nucleotide Archive under the BioProject accession PRJEB20357, individual isolate accession numbers are shown in Table S5.

## Acknowledgements

SS acknowledges the support of the NIHR Imperial Biomedical Research Centre (BRC). EJ is a Rosetrees/Stoneygate 2017 Imperial College Research Fellow (M683). CEC is supported by a personal NIHR Career Development Fellowship (NIHR-2016-090-015). Imperial College London is grateful for the support from the North West London NIHR Applied Research Collaboration. The funders had no role in study design or writing.

## Funding

The research was funded by the National Institute for Health Research Health Protection Research Unit (NIHR HPRU) in Healthcare Associated Infections and Antimicrobial Resistance at Imperial College London in partnership with Public Health England (HPRU-2012-10047). This report is independent research funded by the National Institute for Health Research. The views expressed in this publication are those of the author(s) and not necessarily those of the NHS, the National Institute for Health Research, the Department of Health and Social Care or Public Health England. NW and MJE were also supported by PHE and the Joint Programming Initiative on Antimicrobial Resistance (JPIAMR) and the Medical Research Council (MRC) as part of the ‘ST131TS Consortium’ under grant code MR/R002843/1. The funding sources had no role in study design, data collection, analysis, or decision to submit the manuscript for publication.

## Author contributions

EJ, ME, NW and SS contributed to the study design. KH, OB, EJ, SS and CC verification of methods and data analysis. EJ performed bioinformatics analysis. OB, KH did statistical analysis. OB, KH, MM, FR, MP, JP and EJ contributed to data collection and preparation of samples for whole genome sequencing. OB and SS wrote the ethics application and got the approval. EJ, KH and SS took the lead in writing the manuscript. All authors provided critical feedback of the results and review of the manuscript.

## Conflict of interests

JP is a paid consultant to Next Gen Diagnostics Llc. MJE and NW are members of PHE’s Antimicrobial Resistance and Healthcare Associated Infections Reference Unit, which has received financial support for conference attendance, lectures, research projects, or contracted evaluations from numerous sources, including Accelerate Diagnostics, Achaogen Inc, Allecra Therapeutics, Amplex, AstraZeneca UK Ltd, AusDiagnostics, Basilea Pharmaceutica, Becton Dickinson Diagnostics, bioMérieux, Bio-Rad Laboratories, British Society for Antimicrobial Chemotherapy, Cepheid, Check-Points B.V., Cubist Pharmaceuticals, Department of Health, Enigma Diagnostics, the European Centre for Disease Prevention and Control, Food Standards Agency, GenePOC, GlaxoSmithKline Services Ltd, Helperby Therapeutics, Henry Stewart Talks, International Health Management Associates Ltd, Innovate UK, Kalidex Pharmaceuticals, Melinta Therapeutics, Merck Sharpe and Dohme, Meiji Seika Pharma Co Ltd, Mobidiag, Momentum Biosciences Ltd, Neem Biotech, NIHR, Nordic Pharma Ltd, Norgine Pharmaceuticals, Paratek, Rabiotics Rx, Rempex Pharmaceuticals Ltd, Roche, Rokitan Ltd, Smith and Nephew UK Ltd, Shionogi and Co Ltd, Tetraphase Pharmaceuticals, Trius Therapeutics, VenatoRx Pharmaceuticals, Wockhardt Ltd, and the World Health Organization. All other authors have no competing interest to declare.

## Notes

### Competing Interest Statement

JP is a paid consultant to Next Gen Diagnostics Llc. MJE and NW are members of PHEs Antimicrobial Resistance and Healthcare Associated Infections Reference Unit, which has received financial support for conference attendance, lectures, research projects, or contracted evaluations from numerous sources, including Accelerate Diagnostics, Achaogen Inc, Allecra Therapeutics, Amplex, AstraZeneca UK Ltd, AusDiagnostics, Basilea Pharmaceutica, Becton Dickinson Diagnostics, bioMerieux, Bio-Rad Laboratories, British Society for Antimicrobial Chemotherapy, Cepheid, Check-Points B.V., Cubist Pharmaceuticals, Department of Health, Enigma Diagnostics, the European Centre for Disease Prevention and Control, Food Standards Agency, GenePOC, GlaxoSmithKline Services Ltd, Helperby Therapeutics, Henry Stewart Talks, International Health Management Associates Ltd, Innovate UK, Kalidex Pharmaceuticals, Melinta Therapeutics, Merck Sharpe and Dohme, Meiji Seika Pharma Co Ltd, Mobidiag, Momentum Biosciences Ltd, Neem Biotech, NIHR, Nordic Pharma Ltd, Norgine Pharmaceuticals, Paratek, Rabiotics Rx, Rempex Pharmaceuticals Ltd, Roche, Rokitan Ltd, Smith and Nephew UK Ltd, Shionogi and Co Ltd, Tetraphase Pharmaceuticals, Trius Therapeutics, VenatoRx Pharmaceuticals, Wockhardt Ltd, and the World Health Organization. All other authors have no competing interest to declare.

## References

[1] Wilson J, Elgohari S, Livermore DM, Cookson B, Johnson A, Lamagni T, et al. Trends among pathogens reported as causing bacteraemia in England, 2004-2008. Clin Microbiol Infect 2011;17. https://doi.org/10.1111/j.1469-0691.2010.03262.x.

[2] Public Health England. Laboratory surveillance of Escherichia coli bacteraemia in England, Wales and Northern Ireland□: 2018. Health Protention Report, Volume 13 Number 37, 18 October 2019.

[3] Vihta KD, Stoesser N, Llewelyn MJ, Quan TP, Davies T, Fawcett NJ, et al. Trends over time in Escherichia coli bloodstream infections, urinary tract infections, and antibiotic susceptibilities in Oxfordshire, UK, 1998–2016: a study of electronic health records. Lancet Infect Dis 2018. https://doi.org/10.1016/S1473-3099(18)30353-0.

[4] Bhattacharya A, Nsonwu O, Johnson AP, Hope R. Estimating the incidence and 30-day all-cause mortality rate of Escherichia coli bacteraemia in England by 2020/21. J Hosp Infect 2018;98. https://doi.org/10.1016/j.jhin.2017.09.021.

[5] Naylor NR, Pouwels KB, Hope R, Green N, Henderson KL, Knight GM, et al. The health and cost burden of antibiotic resistant and susceptible Escherichia coli bacteraemia in the English hospital setting: A national retrospective cohort study. PLoS One 2019. https://doi.org/10.1371/journal.pone.0221944.

[6] Lillie PJ, Johnson G, Ivan M, Barlow GD, Moss PJ. Escherichia coli bloodstream infection outcomes and preventability: a six-month prospective observational study. J Hosp Infect 2019;103. https://doi.org/10.1016/j.jhin.2019.05.007.

[7] Blandy O, Honeyford K, Gharbi M, Thomas A, Ramzan F, Ellington MJ, et al. Factors that impact on the burden of Escherichia coli bacteraemia: multivariable regression analysis of 2011–2015 data from West London. J Hosp Infect 2019. https://doi.org/10.1016/j.jhin.2018.10.024.

[8] Wilson J. Applying Pareto analysis to reducing Escherichia coli bloodstream infections. J Infect Prev 2018;19. https://doi.org/10.1177/1757177418795298.

[9] Otter JA, Galletly TJ, Davies F, Hitchcock J, Gilchrist MJ, Dyakova E, et al. Planning to halve Gram-negative bloodstream infection: getting to grips with healthcare-associated Escherichia coli bloodstream infection sources. J Hosp Infect 2019. https://doi.org/10.1016/j.jhin.2018.07.033.

[10] Köhler CD, Dobrindt U. What defines extraintestinal pathogenic Escherichia coli? Int J Med Microbiol 2011. https://doi.org/10.1016/j.ijmm.2011.09.006.

[11] Day MJ, Doumith M, Abernethy J, Hope R, Reynolds R, Wain J, et al. Population structure of Escherichia coli causing bacteraemia in the UK and Ireland between 2001 and 2010. J Antimicrob Chemother 2016. https://doi.org/10.1093/jac/dkw145.

[12] Banerjee R, Johnson JR. A new clone sweeps clean: The enigmatic emergence of Escherichia coli sequence type 131. Antimicrob Agents Chemother 2014. https://doi.org/10.1128/AAC.02824-14.

[13] Stoesser N, Sheppard AE, Pankhurst L, de Maio N, Moore CE, Sebra R, et al. Evolutionary history of the global emergence of the Escherichia coli epidemic clone ST131. MBio 2016. https://doi.org/10.1128/mBio.02162-15.

[14] Price LB, Johnson JR, Aziz M, Clabots C, Johnston B, Tchesnokova V, et al. The epidemic of extended-spectrum-β-lactamase-producing Escherichia coli ST131 is driven by a single highly pathogenic subclone, H30-Rx. MBio 2013. https://doi.org/10.1128/mBio.00377-13.

[15] Kallonen T, Brodrick HJ, Harris SR, Corander J, Brown NM, Martin V, et al. Systematic longitudinal survey of invasive Escherichia coli in England demonstrates a stable population structure only transiently disturbed by the emergence of ST131. Genome Res 2017. https://doi.org/10.1101/gr.216606.116.

[16] Johnson JR, Thuras P, Johnston BD, Weissman SJ, Limaye AP, Riddell K, et al. The Pandemic H30 Subclone of Escherichia coli Sequence Type 131 Is Associated with Persistent Infections and Adverse Outcomes Independent from Its Multidrug Resistance and Associations with Compromised Hosts. Clin Infect Dis 2016. https://doi.org/10.1093/cid/ciw193.

[17] Davies J, Johnson AP, Hope R. Identifying hospital-onset Escherichia coli bacteraemia cases from English mandatory surveillance: the case for applying a two-day post-admission rule. J Hosp Infect 2017. https://doi.org/10.1016/j.jhin.2017.06.031.

[18] Van Walraven C, Austin PC, Jennings A, Quan H, Forster AJ. A modification of the elixhauser comorbidity measures into a point system for hospital death using administrative data. Med Care 2009;47:626–33. https://doi.org/10.1097/MLR.0b013e31819432e5.

[19] Firth D. Bias Reduction of Maximum Likelihood Estimates. Biometrika 1993. https://doi.org/10.2307/2336755.

[20] Van Smeden M, De Groot JAH, Moons KGM, Collins GS, Altman DG, Eijkemans MJC, et al. No rationale for 1 variable per 10 events criterion for binary logistic regression analysis. BMC Med Res Methodol 2016. https://doi.org/10.1186/s12874-016-0267-3.

[21] Ben Zakour NL, Alsheikh-Hussain AS, Ashcroft MM, Khanh Nhu NT, Roberts LW, Stanton-Cook M, et al. Sequential acquisition of virulence and fluoroquinolone resistance has shaped the evolution of Escherichia coli ST131. MBio 2016. https://doi.org/10.1128/mBio.00347-16.

[22] Baltas I, Stockdale T, Tausan M, Kashif A, Anwar J, Anvar J, et al. Impact of antibiotic timing on mortality from Gram-negative bacteraemia in an English district general hospital: the importance of getting it right every time. J Antimicrob Chemother 2021;76. https://doi.org/10.1093/jac/dkaa478.

[23] Evans RN, Pike K, Rogers CA, Reynolds R, Stoddart M, Howe R, et al. Modifiable healthcare factors affecting 28-day survival in bloodstream infection: A prospective cohort study. BMC Infect Dis 2020;20. https://doi.org/10.1186/s12879-020-05262-6.

[24] Goswami C, Fox S, Holden M, Connor M, Leanord A, Evans TJ. Genetic analysis of invasive escherichia coli in scotland reveals determinants of healthcare-associated versus communityacquired infections. Microb Genomics 2018;4. https://doi.org/10.1099/mgen.0.000190.

[25] Trotter CL, Chandra M, Cano R, Larrauri A, Ramsay ME, Brehony C, et al. A surveillance network for meningococcal disease in Europe. FEMS Microbiol Rev 2007;31. https://doi.org/10.1111/j.1574-6976.2006.00060.x.

[26] Olesen B, Frimodt-Møller J, Leihof RF, Struve C, Johnston B, Hansen DS, et al. Temporal trends in antimicrobial resistance and virulence-associated traits within the Escherichia coli sequence type 131 clonal group and its H30 and H30-Rx Subclones, 1968 to 2012. Antimicrob Agents Chemother 2014;58. https://doi.org/10.1128/AAC.03679-14.

[27] Salas-Mera D, Sainz T, Gómez-Gil Mira MR, Méndez-Echevarría A. Gentamicin resistant E. coli as a cause of urinary tract infections in children. Enferm Infecc Microbiol Clin 2017;35. https://doi.org/10.1016/j.eimc.2016.11.001.

[28] Vidal L, Gafter-Gvili A, Borok S, Fraser A, Leibovici L, Paul M. Efficacy and safety of aminoglycoside monotherapy: Systematic review and meta-analysis of randomized controlled trials. J Antimicrob Chemother 2007;60. https://doi.org/10.1093/jac/dkm193.

[29] Shankar-Hari M, Rubenfeld GD. Race, Ethnicity, and Sepsis: Beyond Adjusted Odds Ratios. Crit Care Med 2018. https://doi.org/10.1097/CCM.0000000000003060.

[30] Ayoubkhani D, Nafilyan V, White C, Goldblatt P, Gaughan C, Blackwell L, et al. Ethnic-minority groups in England and Wales-factors associated with the size and timing of elevated COVID-19 mortality: a retrospective cohort study linking census and death records. Int J Epidemiol 2021;49. https://doi.org/10.1093/ije/dyaa208.

