## supplementary material for "Bacterial genotypic and patient risk factors for adverse outcomes in *Escherichia coli* bloodstream infections: a prospective molecular-epidemiological study"

### Supplementary Methods

#### ST131 sub-clade analysis

*E. coli* isolates were grown on MacConkey agar (Oxoid, UK) prior to DNA extraction (Qiasymphony DNA Mini kit, Qiagen). Multiplexed DNA library preparation was conducted according to the Illumina protocol and sequencing was performed on an Illumina HiSeq 2000 (Illumina, USA) with 100-cycle paired-end runs. Genotypes based on multi-locus sequence types (MLST) were determined using SRST2[1] and previously unreported MLST were assigned novel MLST number using *E. coli* database hosted on EnteroBase (http://enterobase.warwick.ac.uk/species/index/ecoli). ST131 genomes were assigned to specific ST131 sub-clades: A, B, C1 and C2 based on previously reported criteria by Price *et al* [2], Petty *et al* [3] and specific SNPs by Ben Zankour *et al* [4]and loss and gain of C2 clade defining CTX-M genes as reported by Kallonen *et al* [5] and Stoesser *et al* [6].

#### Data availability

Whole genome sequencing reads were deposited in European Nucleotide Archive under the BioProject accession PRJEB20357, individual isolate accession numbers are shown in Table S5.

**References**

[1] Inouye M, Dashnow H, Raven LA, Schultz MB, Pope BJ, Tomita T, et al. SRST2: Rapid genomic surveillance for public health and hospital microbiology labs. Genome Med 2014. https://doi.org/10.1186/s13073-014-0090-6.

[2] Price LB, Johnson JR, Aziz M, Clabots C, Johnston B, Tchesnokova V, et al. The epidemic of extended-spectrum-β-lactamase-producing Escherichia coli ST131 is driven by a single highly pathogenic subclone, H30-Rx. MBio 2013. https://doi.org/10.1128/mBio.00377-13.

[3] Petty NK, Zakour NLB, Stanton-Cook M, Skippington E, Totsika M, Forde BM, et al. Global dissemination of a multidrug resistant Escherichia coli clone. Proc Natl Acad Sci U S A 2014. https://doi.org/10.1073/pnas.1322678111.

[4] Ben Zakour NL, Alsheikh-Hussain AS, Ashcroft MM, Khanh Nhu NT, Roberts LW, Stanton-Cook M, et al. Sequential acquisition of virulence and fluoroquinolone resistance has shaped the evolution of Escherichia coli ST131. MBio 2016. https://doi.org/10.1128/mBio.00347-16.

[5] Kallonen T, Brodrick HJ, Harris SR, Corander J, Brown NM, Martin V, et al. Systematic longitudinal survey of invasive Escherichia coli in England demonstrates a stable population structure only transiently disturbed by the emergence of ST131. Genome Res 2017. https://doi.org/10.1101/gr.216606.116.

[6] Stoesser N, Sheppard AE, Pankhurst L, de Maio N, Moore CE, Sebra R, et al. Evolutionary history of the global emergence of the Escherichia coli epidemic clone ST131. MBio 2016. https://doi.org/10.1128/mBio.02162-15.

**Supplementary Figures**

**Supplementary Figure S1. Flowchart of *E. coli* bacteraemia cases included in the analyses in this study.** This flowchart summarises the data available for each of the analyses described in the study.

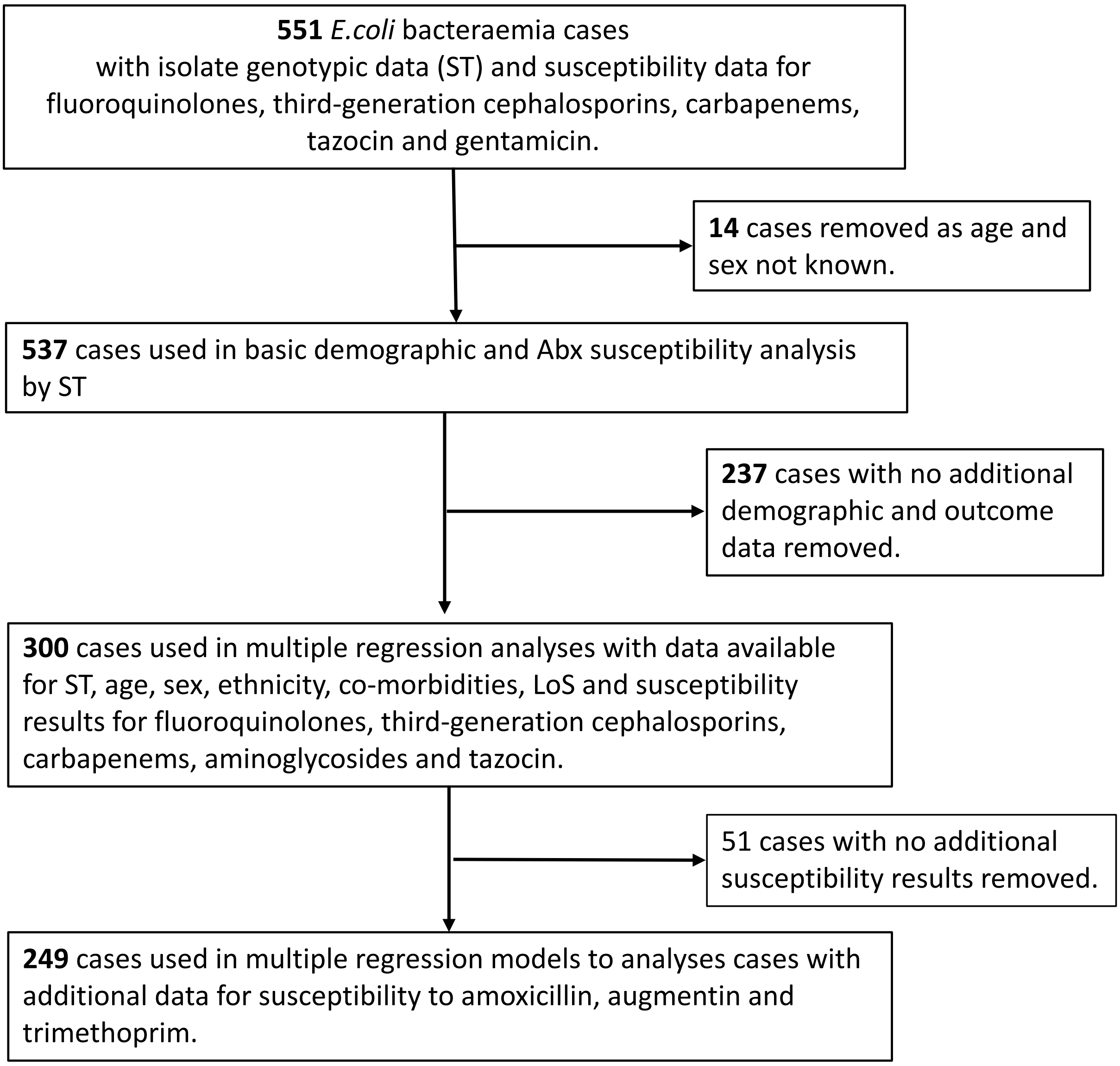

### Supplementary Tables

**Table S1. Summary of patients' characteristics in association with sequence type of *E. coli* causing bacteraemia.** A total of 537 patients had age and gender information available for comparison.

| | ST131-C2 | ST131-other | ST69 | ST73 | ST95 | Other STs | Total |
|---|---|---|---|---|---|---|---|
| **Male** | | | | | | | |
| **< 65** | 11 (18·3%) | 9 (16·4%) | 11 (21·6%) | 13 (16·9%) | 6 (13·3%) | 37 (14·9%) | 87 (16·2%) |
| **≥ 65** | 21 (35·0%) | 15 (27·2%) | 4 (7·8%) | 20 (26·0%) | 6 (13·3%) | 80 (32·1%) | 146 (27·2%) |
| **Total** | 32 (53·3%) | 24 (43·6%) | 15 (29·4%) | 33 (42·9%) | 12 (26·7%) | 117 (47·0%) | 233 (43·4%) |
| **Female** | | | | | | | |
| **< 65** | 5 (8·3%) | 12 (21·9%) | 17 (33·3%) | 15 (19·5%) | 15 (33·3%) | 65 (26·0%) | 129 (24·0%) |
| **≥ 65** | 23 (38·4%) | 19 (34·5%) | 19 (37·3%) | 29 (37·7%) | 18 (37·8%) | 67 (26·8%) | 175 (32·6%) |
| **Total** | 28 (46·7%) | 31 (56·4%) | 36 (70·6%) | 44 (57·1%) | 33 (73·3%) | 132 (53·0%) | 304 (56·6%) |
| **All** | | | | | | | |
| **< 65** | 16 (26·6%) | 21 (38·2%) | 28 (54·9%) | 28 (36·4%) | 21 (46·6%) | 102 (40·9%) | 216 (40·2%) |
| **≥ 65** | 44 (73·4%) | 34 (61·7%) | 23 (45·1%) | 49 (63·7%) | 24 (51·1%) | 147 (59·0%) | 321 (59·8%) |
| **Total** | 60 (11·2%) | 55 (10·2%) | 51 (9·5%) | 77 (14·3%) | 45 (8·4%) | 249 (46·4%) | 537 (100·0%) |

ST131-C2 indicates isolates that were assigned to ST131 clade 2, ST131- other include all non-clade 2 ST131 isolates.

**Table S2. Summary of reported antimicrobial non-susceptibility in association with bacteraemia-causing *E. coli* sequence type.**

| Antibiotic | ST131-C2 (n=60) | ST131-other (n=55) | ST69 (n=51) | ST73 (n=77) | ST95 (n=45) | Other STs (n=249) | Total (n=537) | p-value[1] |
|---|---|---|---|---|---|---|---|---|
| **Fluoroquinolones** | 43 (71·7%) | 28 (50·9%) | 4 (7·8%) | 8 (10·4%) | 3 (6·7%) | 57 (22·9%) | 143 (26·7%) | <0·0001 |
| **Third-generation cephalosporins** | 34 (56·7%) | 15 (27·3%) | 4 (7·8%) | 10 (13·0%) | 2 (4·4%) | 41 (16·5%) | 106 (19·7%) | <0·0001 |
| **Carbapenems** | 0 (0%) | 0 (0%) | 0 (0%) | 0 (0%) | 0 (0%) | 1 (0·4%) | 1 (0·2%) | n/a |
| **Gentamicin** | 21 (65·0%) | 15 (27·3%) | 5 (9·8%) | 7 (9·1%) | 3 (6·7%) | 30 (12 %) | 81 (15·1%) | <0·0001 |
| **Tazocin** | 17 (28·3%) | 4 (7·1%) | 4 (7·8%) | 12 (15·6%) | 0 (0%) | 28 (11·2%) | 65 (12·1%) | 0·0002 |
| **MDR** | 20 (32·8%) | 2 (3·6%) | 0 (0%) | 5 (6·3%) | 2 (4·4%) | 17 (6·6%) | 84 (15·2%) | 0·0005 |
| **Other antibiotics*** | n=27 | n=23 | n=29 | n=33 | n=21 | n=120 | n=249 | |
| **Amoxicillin** | 23 (85·2%) | 18 (78·3%) | 18 (62·1%) | 20 (60·1%) | 11 (52·4%) | 69 (57·5%) | 159 (63·9%) | 0·059 |
| **Augmentin** | 18 (67%) | 11 (47·8%) | 4 (13·8%) | 5 (15·2%) | 4 (19·0%) | 34 (28·3%) | 76 (30·5%) | <0·0001 |
| **Trimethoprim** | 22 (81·5%) | 13 (56·5%) | 14 (48·3%) | 7 (21·2%) | 5 (23·8%) | 50 (41·7%) | 111 (44·6%) | <0·0001 |

[1]p-values are from Chi-squared tests

*antibiotic susceptibility testing results for amoxicillin, augmentin and trimethoprim were available only for 249 out of 537 patient isolates tested. MDR – multi-drug resistance, this included isolates which were non-susceptible to all three classes of the antimicrobial agents (ciprofloxacin, third-generation cephalosporins or gentamicin).

**Table S3. Multiple regression modelling of association between *E. coli* genotype, patient characteristics, antibiotic resistance and mortality within 90 days.** Detailed clinical data and antibiotic susceptibility testing results for augmentin, amoxicillin and tazocin from Local NHS Trust was available for 249 *E. coli* bacteraemia cases.

| | **ST Group – unadjusted** | **Model 1: ST Group adjusted for patient characteristics** | **Model 2: Model 1 + augmentin non-susceptibility** | **Model 3: Model 1 + amoxicillin non-susceptibility** | **Model 4: Model 1 + tazocin non-susceptibility** |
|---|---|---|---|---|---|
| ***E. coli* genotype** | | | | | |
| **Other STs[R]** | | | | | |
| **ST131-C2** | 0·79 (0·23, 2·23) | 0·85 (0·21, 2·98) | 0·74 (0·17, 2·82) | 0·76 (0·18, 2·76) | 0·82 (0·20, 3·02) |
| **ST131** | 2·28 (0·85, 5·81) | 2·73 (0·87, 8·45) | 2·58 (0·82, 8·05) | 2·57 (0·82, 8·00) | 2·70 (0·87, 8·34) |
| **ST69** | 0·38 (0·07, 1·27) | 0·32 (0·03, 1·60) | 0·34 (0·03, 1·70) | 0·33 (0·03, 1·66) | 0·33 (0·03, 1·64) |
| **ST73** | 0·48 (0·12, 1·41) | 0·66 (0·15, 2·31) | 0·67 (0·15, 2·36) | 0·64 (0·14, 2·26) | 0·66 (0·15, 2·33) |
| **ST95** | 0·53 (0·10, 1·84) | 0·67 (0·11, 2·73) | 0·64 (0·11, 2·67) | 0·65 (0·11, 2·68) | 0·67 (0·11, 2·73) |
| **Patient's age** | | | | | |
| **<65[R]** | | | | | |
| **65-74** | | 0·81 (0·23, 2·69) | 0·82 (0·23, 2·73) | 0·84 (0·24, 2·82) | 0·81 (0·23, 2·68) |
| **75-84** | | 1·39 (0·50, 3·90) | 1·39 (0·50, 3·86) | 1·36 (0·49, 3·82) | 1·40 (0·50, 3·91) |
| **85 plus** | | 1·81 (0·47, 6·61) | 1·85 (0·48, 6·90) | 1·98 (0·51, 7·52) | 1·81 (0·48, 6·64) |
| **Patient's sex** | | | | | |
| **Male[R]** | | | | | |
| **Female** | | 0·84 (0·35, 1·97) | 0·84 (0·36, 1·99) | 0·85 (0·36, 1·99) | 0·84 (0·35, 1·98) |
| **Patient's ethnicity** | | | | | |
| **White[R]** | | | | | |

| | | | | | |
|---|---|---|---|---|---|
| **Asian** | | 1·34 (0·39, 4·27) | 1·25 (0·36, 4·05) | 1·16 (0·32, 3·93) | 1·29 (0·36, 4·33) |
| **Black** | | 3·63 (1·16, 11·44) | 3·56 (1·15, 11·09)* | 3·58 (1·15, 11·18)* | 3·51 (1·09, 11·39)* |
| **Other** | | 1·68 (0·44, 6·17) | 1·64 (0·43, 6·00) | 1·61 (0·41, 5·95) | 1·66 (0·43, 6·11) |
| **Not stated** | | 1·39 (0·31, 5·22) | 1·33 (0·29, 5·01) | 1·31 (0·28, 4·96) | 1·36 (0·30, 5·17) |
| **Co-morbidity score** | | | | | |
| **<0[R]** | | | | | |
| **0 to 5** | | 1·30 (0·19, 9·08) | 1·32 (0·19, 9·22) | 1·30 (0·19, 9·11) | 1·30 (0·19, 9·00) |
| **6 to 13** | | 3·93 (1·03, 21·79)* | 3·95 (1·03, 22·06)* | 4·07 (1·05, 22·93)* | 3·86 (1·01, 21·30)* |
| **14 or greater** | | 15·71 (4·46, 85·27)*** | 15·33 (4·34, 83·92)*** | 16·19 (4·54, 90·12)*** | 15·31 (4·38, 82·57) |
| **Onset of infection** | | | | | |
| **Community onset[R]** | | | | | |
| **Hospital onset** | | 0·96 (0·39, 2·25) | 0·97 (0·40, 2·28) | 0·95 (0·39, 2·23) | 0·96 (0·39, 2·25) |
| **Other** | | | | | |
| **Antibiotic non-susceptibility** | | | 1·29 (0·54, 3·03) | 1·37 (0·57, 3·41) | 1·08 (0·46, 2·52) |

Odd ratios are presented with 95% Confidence Intervals; p-values were calculated using Chi square; * p-value of <0·05; ***p-value of <0·001;

[R] group used as reference for the model.

**Table S4. Multiple regression modelling of association between *E. coli* genotype, patient characteristics, antibiotic resistance and dichotomised length of stay.** Detailed clinical data and antibiotic susceptibility testing results for augmentin, amoxicillin and tazocin from Local NHS Trust was available for 249 *E. coli* bacteraemia cases.

| | **ST Group – unadjusted** | **Model 1:** **ST Group adjusted for patient characteristics** | **Model 2:** **Model 1 + augmentin non-susceptibility** | **Model 3:** **Model 1 + amoxicillin non-susceptibility** | **Model 4:** **Model 1 + tazocin non-susceptibility** |
|---|---|---|---|---|---|
| ***E. coli* genotype** | | | | | |
| **Other STs$^{R}$** | | | | | |
| **ST131-C2** | 9·46 (2·33, 86·65)*** | 6·62 (1·50, 63·10) ** | 6·48 (1·44, 62·25)* | 5·53 (1·24, 53·05)* | 6·25 (1·39, 60·00)* |
| **ST131** | 1·18 (0·47, 3·18) | 0·84 (0·31, 2·42) | 0·83 (0·30, 2·42) | 0·76 (0·28, 2·21) | 0·82 (0·30, 2·36) |
| **ST69** | 1·77 (0·71, 4·95) | 3·07 (1·12, 9·37)* | 3·08 (1·12, 9·51)* | 2·92 (1·07, 8·92)* | 3·00 (1·10, 9·16)* |
| **ST73** | 0·92 (0·42, 2·08) | 0·85 (0·35, 2·09) | 0·86 (0·35, 2·11) | 0·82 (0·34, 2·02) | 0·88 (0·36, 2·17) |
| **ST95** | 1·04 (0·41, 2·83) | 1·30 (0·45, 3·96) | 1·30 (0·46, 3·97) | 1·30 (0·45, 3·99) | 1·33 (0·46, 4·07) |
| **Patient's age** | | | | | |
| **<65$^{R}$** | | | | | |
| **65-74** | | 1·41 (0·65, 3·13) | 1·41 (0·65, 3·11) | 1·53 (0·70, 3·41) | 1·43 (0·66, 3·16) |
| **75-84** | | 1·79 (0·77, 4·35) | 1·78 (0·77, 4·32) | 1·84 (0·79, 4·46) | 1·84 (0·79, 4·50) |
| **85 plus** | | 1·61 (0·59, 4·73) | 1·61 (0·59, 4·75) | 1·91 (0·67, 5·84) | 1·67 (0·60, 4·96) |
| **Patient's sex** | | | | | |
| **Male$^{R}$** | | | | | |
| **Female** | | 0·28 (0·14, 0·55)*** | 0·28 (0·14, 0·55)*** | 0·30 (0·15, 0·59)*** | 0·29 (0·14, 0·57)*** |

| | | | | | |
|---|---|---|---|---|---|
| **Ethnicity** | | | | | |
| **White**[R] | | | | | |
| **Asian** | | 1·48 (0·56, 4·23) | 1·47 (0·56, 4·20) | 1·40 (0·52, 3·99) | 1·44 (0·54, 4·11) |
| **Black** | | 0·78 (0·26, 2·43) | 0·78 (0·26, 2·44) | 0·80 (0·27, 2·53) | 0·74 (0·24, 2·33) |
| **Other** | | 0·69 (0·28, 1·75) | 0·69 (0·28, 1·76) | 0·72 (0·29, 1·83) | 0·68 (0·27, 1·73) |
| **Not stated** | | 0·75 (0·30, 1·88) | 0·75 (0·30, 1·88) | 0·75 (0·30, 1·91) | 0·74 (0·30, 1·86) |
| **Co-morbidity score** | | | | | |
| **<0**[R] | | | | | |
| **0 to 5** | | 1·82 (0·79, 4·26) | 1·81 (0·79, 4·24) | 1·76 (0·77, 4·16) | 1·72 (0·74, 4·10) |
| **6 to 13** | | 2·81 (1·23, 6·58)* | 2·79 (1·23, 6·54)* | 2·85 (1·25, 6·70)* | 2·73 (1·19, 6·41)* |
| **14 or greater** | | 2·07 (0·84, 5·23) | 2·06 (0·84, 5·20) | 2·10 (0·86, 5·30) | 2·02 (0·82, 5·10) |
| **Onset of infection** | | | | | |
| **Community onset**[R] | | | | | |
| **Hospital onset** | | 2·36 (1·05, 5·71)* | 2·35 (1·05, 5·69)* | 2·38 (1·05, 5·78)* | 2·32 (1·03, 5·60) |
| **Other** | | | | | |
| **Antibiotic non-susceptibility** | | NA | 1·03 (0·50, 2·17) | 1·53 (0·81, 2·91) | 1·20 (0·62, 2·34) |

Odd ratios are presented with 95% Confidence Intervals; p-values were calculated using Chi square; * p-value of <0·05; ***p-value of <0·001;

[R] group used as reference for the model.

**Table 5. Accession numbers for the *E. coli* blood isolates sequenced as part of this study.**

| HPRU_ID | accession no. | MLST (ST) |
|---|---|---|
| HPRU-EC-0002 | ERS1703537 | 7519 |
| HPRU-EC-0003 | ERS1703538 | 131 |
| HPRU-EC-0004 | ERS1703539 | 73 |
| HPRU-EC-0005 | ERS1703540 | 73 |
| HPRU-EC-0006 | ERS1703529 | 131 |
| HPRU-EC-0007 | ERS1703530 | 73 |
| HPRU-EC-0008 | ERS1703531 | 73 |
| HPRU-EC-0010 | ERS1703533 | 38 |
| HPRU-EC-0011 | ERS1703534 | 354 |
| HPRU-EC-0012 | ERS1703535 | 73 |
| HPRU-EC-0014 | ERS1703541 | 95 |
| HPRU-EC-0015 | ERS1703542 | 393 |
| HPRU-EC-0016 | ERS1703543 | 104 |
| HPRU-EC-0017 | ERS1703544 | 95 |
| HPRU-EC-0018 | ERS1703545 | 648 |
| HPRU-EC-0019 | ERS1703546 | 7520 |
| HPRU-EC-0020 | ERS1703547 | 38 |
| HPRU-EC-0022 | ERS1703549 | 131 |
| HPRU-EC-0023 | ERS1703550 | 69 |
| HPRU-EC-0024 | ERS1703551 | 131 |
| HPRU-EC-0026 | ERS1703553 | 484 |
| HPRU-EC-0027 | ERS1703554 | 73 |
| HPRU-EC-0028 | ERS1703555 | 62 |
| HPRU-EC-0029 | ERS1703556 | 73 |
| HPRU-EC-0030 | ERS1703557 | 131 |
| HPRU-EC-0031 | ERS1703558 | 140 |
| HPRU-EC-0032 | ERS1703559 | 38 |
| HPRU-EC-0033 | ERS1703560 | 62 |
| HPRU-EC-0034 | ERS1703561 | 2967 |
| HPRU-EC-0035 | ERS1703562 | 3312 |
| HPRU-EC-0036 | ERS1703563 | 1262 |
| HPRU-EC-0037 | ERS1703564 | 131 |
| HPRU-EC-0038 | ERS1703565 | 73 |
| HPRU-EC-0039 | ERS1703566 | 69 |
| HPRU-EC-0042 | ERS1703567 | 405 |
| HPRU-EC-0043 | ERS1703568 | 12 |

| | | |
|---|---|---|
| HPRU-EC-0047 | ERS1703569 | 69 |
| HPRU-EC-0048 | ERS1703570 | 73 |
| HPRU-EC-0049 | ERS1703571 | 73 |
| HPRU-EC-0050 | ERS1703572 | 550 |
| HPRU-EC-0051 | ERS1703573 | 69 |
| HPRU-EC-0052 | ERS1703574 | 95 |
| HPRU-EC-0055 | ERS1703575 | 69 |
| HPRU-EC-0056 | ERS1703576 | 131 |
| HPRU-EC-0058 | ERS1703577 | 73 |
| HPRU-EC-0060 | ERS1703578 | 131 |
| HPRU-EC-0061 | ERS1703579 | 95 |
| HPRU-EC-0064 | ERS1703580 | 131 |
| HPRU-EC-0065 | ERS1703581 | 93 |
| HPRU-EC-0067 | ERS1703582 | 131 |
| HPRU-EC-0069 | ERS1703583 | 1316 |
| HPRU-EC-0072 | ERS1703584 | 7521 |
| HPRU-EC-0073 | ERS1703585 | 1316 |
| HPRU-EC-0074 | ERS1703586 | 7521 |
| HPRU-EC-0079 | ERS1703588 | 69 |
| HPRU-EC-0082 | ERS1703589 | 38 |
| HPRU-EC-0084 | ERS1703590 | 95 |
| HPRU-EC-0085 | ERS1703591 | 73 |
| HPRU-EC-0086 | ERS1703592 | 73 |
| HPRU-EC-0088 | ERS1703623 | 12 |
| HPRU-EC-0089 | ERS1703655 | 624 |
| HPRU-EC-0092 | ERS1703658 | 131 |
| HPRU-EC-0093 | ERS1703659 | 624 |
| HPRU-EC-0095 | ERS1703660 | 624 |
| HPRU-EC-0096 | ERS1703661 | 131 |
| HPRU-EC-0097 | ERS1703662 | 12 |
| HPRU-EC-0098 | ERS1703663 | 88 |
| HPRU-EC-0100 | ERS1703664 | 69 |
| HPRU-EC-0101 | ERS1703665 | 73 |
| HPRU-EC-0103 | ERS1703666 | 88 |
| HPRU-EC-0104 | ERS1703667 | 69 |
| HPRU-EC-0107 | ERS1703668 | 95 |
| HPRU-EC-0109 | ERS1703669 | 2015 |
| HPRU-EC-0110 | ERS1703670 | 88 |
| HPRU-EC-0111 | ERS1703671 | 155 |
| HPRU-EC-0113 | ERS1703672 | 62 |
| HPRU-EC-0114 | ERS1703673 | 141 |
| HPRU-EC-0115 | ERS1703674 | 410 |

| | | |
|---|---|---|
| HPRU-EC-0119 | ERS1703675 | 131 |
| HPRU-EC-0128 | ERS1703676 | 73 |
| HPRU-EC-0129 | ERS1703677 | 73 |
| HPRU-EC-0135 | ERS1703678 | 131 |
| HPRU-EC-0136 | ERS1703679 | 4373 |
| HPRU-EC-0137 | ERS1703681 | 14 |
| HPRU-EC-0138 | ERS1703682 | 131 |
| HPRU-EC-0139 | ERS1703683 | 538 |
| HPRU-EC-0142 | ERS1703684 | 95 |
| HPRU-EC-0145 | ERS1703685 | 95 |
| HPRU-EC-0149 | ERS1703686 | 393 |
| HPRU-EC-0153 | ERS1703688 | 929 |
| HPRU-EC-0156 | ERS1703690 | 73 |
| HPRU-EC-0157 | ERS1703691 | 69 |
| HPRU-EC-0158 | ERS1703692 | 69 |
| HPRU-EC-0159 | ERS1703693 | 69 |
| HPRU-EC-0160 | ERS1703694 | 38 |
| HPRU-EC-0164 | ERS1703695 | 95 |
| HPRU-EC-0167 | ERS1703696 | 73 |
| HPRU-EC-0170 | ERS1703698 | 131 |
| HPRU-EC-0172 | ERS1703699 | 69 |
| HPRU-EC-0173 | ERS1703701 | 73 |
| HPRU-EC-0174 | ERS1703702 | 352 |
| HPRU-EC-0176 | ERS1703703 | 10 |
| HPRU-EC-0179 | ERS1703705 | 73 |
| HPRU-EC-0182 | ERS1703707 | 1057 |
| HPRU-EC-0183 | ERS1703708 | 69 |
| HPRU-EC-0184 | ERS1703709 | 372 |
| HPRU-EC-0185 | ERS1703710 | 73 |
| HPRU-EC-0186 | ERS1703711 | 95 |
| HPRU-EC-0187 | ERS1703712 | 73 |
| HPRU-EC-0191 | ERS1703713 | 95 |
| HPRU-EC-0192 | ERS1703714 | 69 |
| HPRU-EC-0193 | ERS1703715 | 93 |
| HPRU-EC-0196 | ERS1703716 | 93 |
| HPRU-EC-0203 | ERS1703718 | 429 |
| HPRU-EC-0208 | ERS1703719 | 1057 |
| HPRU-EC-0209 | ERS1703720 | 10 |
| HPRU-EC-0210 | ERS1703721 | 1057 |
| HPRU-EC-0211 | ERS1703722 | 131 |
| HPRU-EC-0212 | ERS1703723 | 1914 |
| HPRU-EC-0215 | ERS1703724 | 131 |

| | | |
|---|---|---|
| HPRU-EC-0217 | ERS1703725 | 131 |
| HPRU-EC-0218 | ERS1703726 | 69 |
| HPRU-EC-0219 | ERS1703727 | 73 |
| HPRU-EC-0221 | ERS1703728 | 1652 |
| HPRU-EC-0230 | ERS1703729 | 131 |
| HPRU-EC-0231 | ERS1703730 | 12 |
| HPRU-EC-0232 | ERS1703731 | 73 |
| HPRU-EC-0236 | ERS1703733 | 2851 |
| HPRU-EC-0237 | ERS1703734 | 12 |
| HPRU-EC-0239 | ERS1703735 | 14 |
| HPRU-EC-0242 | ERS1703737 | 69 |
| HPRU-EC-0243 | ERS1703740 | 73 |
| HPRU-EC-0244 | ERS1703738 | 131 |
| HPRU-EC-0245 | ERS1703741 | 7523 |
| HPRU-EC-0247 | ERS1703742 | 131 |
| HPRU-EC-0248 | ERS1703743 | 12 |
| HPRU-EC-0250 | ERS1703744 | 131 |
| HPRU-EC-0260 | ERS1703745 | 131 |
| HPRU-EC-0261 | ERS1703746 | 131 |
| HPRU-EC-0266 | ERS1703747 | 95 |
| HPRU-EC-0267 | ERS1703748 | 62 |
| HPRU-EC-0268 | ERS1703749 | 73 |
| HPRU-EC-0269 | ERS1703750 | 6756 |
| HPRU-EC-0270 | ERS1703751 | 69 |
| HPRU-EC-0271 | ERS1703752 | 131 |
| HPRU-EC-0272 | ERS1703753 | 73 |
| HPRU-EC-0273 | ERS1703754 | 73 |
| HPRU-EC-0274 | ERS1703755 | 73 |
| HPRU-EC-0283 | ERS1703756 | 7525 |
| HPRU-EC-0284 | ERS1703757 | 38 |
| HPRU-EC-0286 | ERS1703758 | 127 |
| HPRU-EC-0290 | ERS1703759 | 69 |
| HPRU-EC-0291 | ERS1703760 | 131 |
| HPRU-EC-0292 | ERS1703761 | 131 |
| HPRU-EC-0293 | ERS1703762 | 73 |
| HPRU-EC-0294 | ERS1703763 | 131 |
| HPRU-EC-0296 | ERS1703765 | 69 |
| HPRU-EC-0297 | ERS1703766 | 131 |
| HPRU-EC-0299 | ERS1703767 | 69 |
| HPRU-EC-0300 | ERS1703768 | 2175 |
| HPRU-EC-0301 | ERS1703769 | 38 |
| HPRU-EC-0303 | ERS1703770 | 73 |

| | | |
|---|---|---|
| HPRU-EC-0304 | ERS1703771 | 73 |
| HPRU-EC-0306 | ERS1703773 | 38 |
| HPRU-EC-0308 | ERS1703775 | 69 |
| HPRU-EC-0309 | ERS1703774 | 95 |
| HPRU-EC-0311 | ERS1703776 | 978 |
| HPRU-EC-0313 | ERS1703777 | 117 |
| HPRU-EC-0314 | ERS1703778 | 73 |
| HPRU-EC-0315 | ERS1703781 | 2279 |
| HPRU-EC-0319 | ERS1703782 | 131 |
| HPRU-EC-0321 | ERS1703783 | 117 |
| HPRU-EC-0322 | ERS1703784 | 88 |
| HPRU-EC-0325 | ERS1703785 | 131 |
| HPRU-EC-0327 | ERS1703786 | 95 |
| HPRU-EC-0328 | ERS1703787 | 648 |
| HPRU-EC-0329 | ERS1703788 | 141 |
| HPRU-EC-0330 | ERS1703789 | 131 |
| HPRU-EC-0333 | ERS1703790 | 95 |
| HPRU-EC-0334 | ERS1703791 | 978 |
| HPRU-EC-0335 | ERS1703792 | 1193 |
| HPRU-EC-0337 | ERS1703793 | 131 |
| HPRU-EC-0338 | ERS1703794 | 219 |
| HPRU-EC-0340 | ERS1703797 | 1808 |
| HPRU-EC-0342 | ERS1703798 | 69 |
| HPRU-EC-0344 | ERS1703799 | 73 |
| HPRU-EC-0362 | ERS1703801 | 69 |
| HPRU-EC-0363 | ERS1703802 | 131 |
| HPRU-EC-0364 | ERS1703803 | 131 |
| HPRU-EC-0365 | ERS1703804 | 420 |
| HPRU-EC-0366 | ERS1703805 | 131 |
| HPRU-EC-0367 | ERS1703806 | 62 |
| HPRU-EC-0368 | ERS1703807 | 162 |
| HPRU-EC-0370 | ERS1703808 | 362 |
| HPRU-EC-0371 | ERS1703809 | 537 |
| HPRU-EC-0372 | ERS1703810 | 73 |
| HPRU-EC-0373 | ERS1703811 | 131 |
| HPRU-EC-0375 | ERS1703812 | 131 |
| HPRU-EC-0376 | ERS1703813 | 69 |
| HPRU-EC-0377 | ERS1703814 | 362 |
| HPRU-EC-0379 | ERS1703815 | 73 |
| HPRU-EC-0380 | ERS1703816 | 73 |
| HPRU-EC-0382 | ERS1703817 | 131 |
| HPRU-EC-0384 | ERS1703818 | 59 |

| | | |
|---|---|---|
| HPRU-EC-0386 | ERS1703819 | 131 |
| HPRU-EC-0387 | ERS1703820 | 131 |
| HPRU-EC-0389 | ERS1703821 | 69 |
| HPRU-EC-0390 | ERS1703822 | 1064 |
| HPRU-EC-0391 | ERS1703823 | 12 |
| HPRU-EC-0392 | ERS1703824 | 95 |
| HPRU-EC-0398 | ERS1703825 | 73 |
| HPRU-EC-0407 | ERS1703826 | 117 |
| HPRU-EC-0408 | ERS1703827 | 131 |
| HPRU-EC-0409 | ERS1703828 | 95 |
| HPRU-EC-0412 | ERS1703830 | 646 |
| HPRU-EC-0414 | ERS1703831 | 12 |
| HPRU-EC-0416 | ERS1703832 | 73 |
| HPRU-EC-0422 | ERS1703834 | 73 |
| HPRU-EC-0423 | ERS1703835 | 648 |
| HPRU-EC-0424 | ERS1703836 | 1249 |
| HPRU-EC-0427 | ERS1703837 | 59 |
| HPRU-EC-0434 | ERS1703838 | 73 |
| HPRU-EC-0436 | ERS1703839 | 12 |
| HPRU-EC-0439 | ERS1703840 | 131 |
| HPRU-EC-0442 | ERS1703841 | 131 |
| HPRU-EC-0443 | ERS1703842 | 421 |
| HPRU-EC-0444 | ERS1703843 | 95 |
| HPRU-EC-0446 | ERS1703844 | 12 |
| HPRU-EC-0449 | ERS1703845 | 69 |
| HPRU-EC-0450 | ERS1703846 | 131 |
| HPRU-EC-0452 | ERS1703847 | 131 |
| HPRU-EC-0454 | ERS1703848 | 648 |
| HPRU-EC-0458 | ERS1703870 | 69 |
| HPRU-EC-0462 | ERS1703872 | 12 |
| HPRU-EC-0466 | ERS1703850 | 131 |
| HPRU-EC-0467 | ERS1703851 | 648 |
| HPRU-EC-0470 | ERS1703852 | 7527 |
| HPRU-EC-0477 | ERS1703853 | 131 |
| HPRU-EC-0480 | ERS1703854 | 59 |
| HPRU-EC-0481 | ERS1703855 | 58 |
| HPRU-EC-0484 | ERS1703856 | 95 |
| HPRU-EC-0485 | ERS1703857 | 349 |
| HPRU-EC-0486 | ERS1703858 | 69 |
| HPRU-EC-0487 | ERS1703859 | 73 |
| HPRU-EC-0491 | ERS1703863 | 131 |
| HPRU-EC-0492 | ERS1703864 | 95 |

| | | |
|---|---|---|
| HPRU-EC-0494 | ERS1703865 | 491 |
| HPRU-EC-0499 | ERS1703866 | 10 |
| HPRU-EC-0504 | ERS1703867 | 10 |
| HPRU-EC-0505 | ERS1703868 | 2491 |
| HPRU-EC-0511 | ERS1703874 | 131 |
| HPRU-EC-0512 | ERS1703875 | 10 |
| HPRU-EC-0513 | ERS1703876 | 73 |
| HPRU-EC-0514 | ERS1703877 | 73 |
| HPRU-EC-0518 | ERS1703880 | 95 |
| HPRU-EC-0520 | ERS1703883 | 10 |
| HPRU-EC-0521 | ERS1703884 | 62 |
| HPRU-EC-0523 | ERS1703885 | 95 |
| HPRU-EC-0525 | ERS1703886 | 357 |
| HPRU-EC-0529 | ERS1703887 | 131 |
| HPRU-EC-0530 | ERS1703888 | 144 |
| HPRU-EC-0531 | ERS1703889 | 131 |
| HPRU-EC-0532 | ERS1703890 | 95 |
| HPRU-EC-0533 | ERS1703891 | 1193 |
| HPRU-EC-0539 | ERS1703894 | 2522 |
| HPRU-EC-0542 | ERS1703895 | 131 |
| HPRU-EC-0544 | ERS1703897 | 2491 |
| HPRU-EC-0545 | ERS1703898 | 101 |
| HPRU-EC-0548 | ERS1703899 | 38 |
| HPRU-EC-0549 | ERS1703900 | 73 |
| HPRU-EC-0550 | ERS1703901 | 117 |
| HPRU-EC-0553 | ERS1703902 | 448 |
| HPRU-EC-0554 | ERS1703904 | 131 |
| HPRU-EC-0555 | ERS1703905 | 131 |
| HPRU-EC-0557 | ERS1703906 | 131 |
| HPRU-EC-0561 | ERS1703907 | 69 |
| HPRU-EC-0562 | ERS1703908 | 131 |
| HPRU-EC-0567 | ERS1703909 | 141 |
| HPRU-EC-0569 | ERS1703910 | 69 |
| HPRU-EC-0570 | ERS1703912 | 69 |
| HPRU-EC-0573 | ERS1703914 | 73 |
| HPRU-EC-0574 | ERS1703915 | 131 |
| HPRU-EC-0576 | ERS1703916 | 761 |
| HPRU-EC-0579 | ERS1703918 | 69 |
| HPRU-EC-0583 | ERS1703919 | 69 |
| HPRU-EC-0586 | ERS1703920 | 636 |
| HPRU-EC-0588 | ERS1703921 | 636 |
| HPRU-EC-0590 | ERS1703922 | 1193 |

| | | |
|---|---|---|
| HPRU-EC-0591 | ERS1703923 | 69 |
| HPRU-EC-0596 | ERS1703924 | 14 |
| HPRU-EC-0597 | ERS1703925 | 1193 |
| HPRU-EC-0601 | ERS1703926 | 7522 |
| HPRU-EC-0604 | ERS1703927 | 95 |
| HPRU-EC-0605 | ERS1703928 | 73 |
| HPRU-EC-0606 | ERS1703929 | 354 |
| HPRU-EC-0611 | ERS1703930 | 69 |
| HPRU-EC-0613 | ERS1703931 | 131 |
| HPRU-EC-0616 | ERS1703932 | 131 |
| HPRU-EC-0623 | ERS1703933 | 58 |
| HPRU-EC-0628 | ERS1703934 | 117 |
| HPRU-EC-0629 | ERS1703935 | 131 |
| HPRU-EC-0630 | ERS1703936 | 12 |
| HPRU-EC-0631 | ERS1703937 | 131 |
| HPRU-EC-0632 | ERS1703938 | 95 |
| HPRU-EC-0634 | ERS1703940 | 80 |
| HPRU-EC-0636 | ERS1703941 | 1193 |
| HPRU-EC-0639 | ERS1703942 | 7516 |
| HPRU-EC-0644 | ERS1703943 | 155 |
| HPRU-EC-0646 | ERS1703944 | 131 |
| HPRU-EC-0652 | ERS1703945 | 95 |
| HPRU-EC-0654 | ERS1703946 | 131 |
| HPRU-EC-0656 | ERS1703947 | 69 |
| HPRU-EC-0657 | ERS1703948 | 404 |
| HPRU-EC-0659 | ERS1703950 | 95 |
| HPRU-EC-0661 | ERS1703951 | 131 |
| HPRU-EC-0662 | ERS1703952 | 95 |
| HPRU-EC-0664 | ERS1703953 | 131 |
| HPRU-EC-0668 | ERS1703955 | 131 |
| HPRU-EC-0670 | ERS1703956 | 428 |
| HPRU-EC-0671 | ERS1703957 | 127 |
| HPRU-EC-0672 | ERS1703958 | 12 |
| HPRU-EC-0673 | ERS1703959 | 73 |
| HPRU-EC-0674 | ERS1703960 | 345 |
| HPRU-EC-0678 | ERS1703962 | 127 |
| HPRU-EC-0679 | ERS1703963 | 2020 |
| HPRU-EC-0681 | ERS1703964 | 131 |
| HPRU-EC-0683 | ERS1703966 | 144 |
| HPRU-EC-0690 | ERS1703967 | 131 |
| HPRU-EC-0691 | ERS1703968 | 131 |
| HPRU-EC-0692 | ERS1703969 | 73 |

| | | |
|---|---|---|
| HPRU-EC-0693 | ERS1703970 | 681 |
| HPRU-EC-0694 | ERS1703971 | 73 |
| HPRU-EC-0698 | ERS1703972 | 38 |
| HPRU-EC-0703 | ERS1703974 | 127 |
| HPRU-EC-0704 | ERS1703975 | 69 |
| HPRU-EC-0706 | ERS1703976 | 73 |
| HPRU-EC-0709 | ERS1703977 | 12 |
| HPRU-EC-0712 | ERS1703981 | 131 |
| HPRU-EC-0716 | ERS1703982 | 14 |
| HPRU-EC-0730 | ERS1703983 | 73 |
| HPRU-EC-0732 | ERS1703984 | 131 |
| HPRU-EC-0736 | ERS1703985 | 1177 |
| HPRU-EC-0738 | ERS1703986 | 69 |
| HPRU-EC-0741 | ERS1703988 | 80 |
| HPRU-EC-0745 | ERS1703989 | 12 |
| HPRU-EC-0747 | ERS1703990 | 73 |
| HPRU-EC-0749 | ERS1703991 | 681 |
| HPRU-EC-0750 | ERS1703993 | 131 |
| HPRU-EC-0752 | ERS1703994 | 636 |
| HPRU-EC-0753 | ERS1703995 | 648 |
| HPRU-EC-0754 | ERS1703996 | 1193 |
| HPRU-EC-0755 | ERS1703997 | 131 |
| HPRU-EC-0756 | ERS1703998 | 131 |
| HPRU-EC-0758 | ERS1703999 | 7524 |
| HPRU-EC-0761 | ERS1704000 | 2741 |
| HPRU-EC-0769 | ERS1704001 | 73 |
| HPRU-EC-0771 | ERS1704002 | 131 |
| HPRU-EC-0777 | ERS1704003 | 73 |
| HPRU-EC-0779 | ERS1704004 | 73 |
| HPRU-EC-0780 | ERS1704005 | 73 |
| HPRU-EC-0781 | ERS1704006 | 69 |
| HPRU-EC-0783 | ERS1704007 | 1859 |
| HPRU-EC-0784 | ERS1704008 | 69 |
| HPRU-EC-0789 | ERS1704009 | 73 |
| HPRU-EC-0792 | ERS1704010 | 58 |
| HPRU-EC-0794 | ERS1704011 | 73 |
| HPRU-EC-0796 | ERS1704012 | 165 |
| HPRU-EC-0797 | ERS1704013 | 80 |
| HPRU-EC-0798 | ERS1704014 | 73 |
| HPRU-EC-0799 | ERS1704015 | 617 |
| HPRU-EC-0804 | ERS1704016 | 73 |
| HPRU-EC-0812 | ERS1704018 | 12 |

| | | |
|---|---|---|
| HPRU-EC-0815 | ERS1704019 | 131 |
| HPRU-EC-0835 | ERS1704020 | 131 |
| HPRU-EC-0836 | ERS1704021 | 224 |
| HPRU-EC-0839 | ERS1704022 | 12 |
| HPRU-EC-0845 | ERS1704023 | 550 |
| HPRU-EC-0846 | ERS1704024 | 95 |
| HPRU-EC-0849 | ERS1704025 | 69 |
| HPRU-EC-0851 | ERS1704026 | 141 |
| HPRU-EC-0852 | ERS1704027 | 12 |
| HPRU-EC-0855 | ERS1704028 | 421 |
| HPRU-EC-0856 | ERS1704029 | 38 |
| HPRU-EC-0858 | ERS1704030 | 131 |
| HPRU-EC-0863 | ERS1704031 | 3489 |
| HPRU-EC-0865 | ERS1704032 | 14 |
| HPRU-EC-0868 | ERS1704033 | 73 |
| HPRU-EC-0870 | ERS1704034 | 73 |
| HPRU-EC-0872 | ERS1704035 | 95 |
| HPRU-EC-0876 | ERS1704036 | 69 |
| HPRU-EC-0877 | ERS1704037 | 1324 |
| HPRU-EC-0878 | ERS1704038 | 95 |
| HPRU-EC-0879 | ERS1704039 | 162 |
| HPRU-EC-0880 | ERS1704040 | 58 |
| HPRU-EC-0884 | ERS1704041 | 131 |
| HPRU-EC-0888 | ERS1704042 | 7526 |
| HPRU-EC-0889 | ERS1704043 | 73 |
| HPRU-EC-0890 | ERS1704044 | 354 |
| HPRU-EC-0892 | ERS1704045 | 569 |
| HPRU-EC-0893 | ERS1704046 | 95 |
| HPRU-EC-0895 | ERS1704047 | 127 |
| HPRU-EC-0896 | ERS1704048 | 73 |
| HPRU-EC-0897 | ERS1704049 | 12 |
| HPRU-EC-0899 | ERS1704051 | 131 |
| HPRU-EC-0902 | ERS1704052 | 7529 |
| HPRU-EC-0904 | ERS1704053 | 62 |
| HPRU-EC-0905 | ERS1704054 | 131 |
| HPRU-EC-0906 | ERS1704055 | 117 |
| HPRU-EC-0907 | ERS1704056 | 73 |
| HPRU-EC-0909 | ERS1704057 | 131 |
| HPRU-EC-0912 | ERS1704058 | 7528 |
| HPRU-EC-0917 | ERS1704059 | 69 |
| HPRU-EC-0919 | ERS1704060 | 501 |
| HPRU-EC-0920 | ERS1704061 | 69 |

| | | |
|---|---|---|
| HPRU-EC-0922 | ERS1704062 | 131 |
| HPRU-EC-0923 | ERS1704063 | 58 |
| HPRU-EC-0924 | ERS1704064 | 131 |
| HPRU-EC-0927 | ERS1704065 | 95 |
| HPRU-EC-0928 | ERS1704066 | 95 |
| HPRU-EC-0929 | ERS1704068 | 131 |
| HPRU-EC-0932 | ERS1704069 | 131 |
| HPRU-EC-0933 | ERS1704070 | 12 |
| HPRU-EC-0935 | ERS1704071 | 131 |
| HPRU-EC-0936 | ERS1704072 | 127 |
| HPRU-EC-0937 | ERS1704073 | 58 |
| HPRU-EC-0938 | ERS1704074 | 95 |
| HPRU-EC-0941 | ERS1704075 | 706 |
| HPRU-EC-0942 | ERS1704076 | 131 |
| HPRU-EC-0944 | ERS1704077 | 95 |
| HPRU-EC-0946 | ERS1704078 | 46 |
| HPRU-EC-0947 | ERS1704079 | 131 |
| HPRU-EC-0948 | ERS1704080 | 372 |
| HPRU-EC-0951 | ERS1704081 | 73 |
| HPRU-EC-0952 | ERS1704082 | 372 |
| HPRU-EC-0953 | ERS1704083 | 372 |
| HPRU-EC-0958 | ERS1704084 | 162 |
| HPRU-EC-0959 | ERS1704086 | 95 |
| HPRU-EC-0963 | ERS1704087 | 131 |
| HPRU-EC-0965 | ERS1704088 | 2473 |
| HPRU-EC-0967 | ERS1704089 | 95 |
| HPRU-EC-0968 | ERS1704090 | 95 |
| HPRU-EC-0971 | ERS1704091 | 537 |
| HPRU-EC-0972 | ERS1704092 | 95 |
| HPRU-EC-0973 | ERS1704093 | 1193 |
| HPRU-EC-0974 | ERS1704094 | 131 |
| HPRU-EC-0975 | ERS1704095 | 127 |
| HPRU-EC-0977 | ERS1704096 | 827 |
| HPRU-EC-0978 | ERS1704097 | 827 |
| HPRU-EC-0979 | ERS1704112 | 404 |
| HPRU-EC-0982 | ERS1704114 | 2448 |
| HPRU-EC-0983 | ERS1704115 | 95 |
| HPRU-EC-0984 | ERS1704116 | 393 |
| HPRU-EC-0985 | ERS1704117 | 69 |
| HPRU-EC-0990 | ERS1704118 | 73 |
| HPRU-EC-0991 | ERS1704119 | 827 |
| HPRU-EC-0993 | ERS1704120 | 83 |

| | | |
|---|---|---|
| HPRU-EC-0995 | ERS1704121 | 127 |
| HPRU-EC-0997 | ERS1704098 | 12 |
| HPRU-EC-1002 | ERS1704099 | 73 |
| HPRU-EC-1003 | ERS1704100 | 73 |
| HPRU-EC-1005 | ERS1704102 | 7514 |
| HPRU-EC-1007 | ERS1704103 | 127 |
| HPRU-EC-1008 | ERS1704104 | 73 |
| HPRU-EC-1009 | ERS1704105 | 998 |
| HPRU-EC-1010 | ERS1704106 | 38 |
| HPRU-EC-1011 | ERS1704107 | 131 |
| HPRU-EC-1016 | ERS1704108 | 10 |
| HPRU-EC-1018 | ERS1704110 | 2279 |
| HPRU-EC-1024 | ERS1704111 | 62 |
| HPRU-EC-1025 | ERS1704122 | 131 |
| HPRU-EC-1026 | ERS1704123 | 127 |
| HPRU-EC-1027 | ERS1704125 | 131 |
| HPRU-EC-1030 | ERS1704126 | 90 |
| HPRU-EC-1032 | ERS1704127 | 131 |
| HPRU-EC-1033 | ERS1704128 | 73 |
| HPRU-EC-1035 | ERS1704129 | 95 |
| HPRU-EC-1037 | ERS1704130 | 131 |
| HPRU-EC-1038 | ERS1704131 | 999 |
| HPRU-EC-1039 | ERS1704132 | 127 |
| HPRU-EC-1040 | ERS1704133 | 167 |
| HPRU-EC-1043 | ERS1704134 | 73 |
| HPRU-EC-1045 | ERS1704135 | 69 |
| HPRU-EC-1050 | ERS1704137 | 69 |
| HPRU-EC-1052 | ERS1704138 | 95 |
| HPRU-EC-1056 | ERS1704139 | 131 |
| HPRU-EC-1057 | ERS1704140 | 1193 |
| HPRU-EC-1059 | ERS1704141 | 404 |
| HPRU-EC-1061 | ERS1704142 | 7513 |
| HPRU-EC-1062 | ERS1704143 | 131 |
| HPRU-EC-1065 | ERS1704144 | 69 |
| HPRU-EC-1068 | ERS1704145 | 404 |
| HPRU-EC-1069 | ERS1704146 | 648 |
| HPRU-EC-1070 | ERS1704147 | 58 |
| HPRU-EC-1072 | ERS1704148 | 131 |
| HPRU-EC-1074 | ERS1704149 | 127 |
| HPRU-EC-1075 | ERS1704150 | 131 |
| HPRU-EC-1076 | ERS1704151 | 144 |
| HPRU-EC-1077 | ERS1704152 | 12 |

| | | |
|---|---|---|
| HPRU-EC-1082 | ERS1704153 | 12 |
| HPRU-EC-1084 | ERS1704154 | 3268 |
| HPRU-EC-1087 | ERS1704155 | 155 |
| HPRU-EC-1088 | ERS1704156 | 14 |
| HPRU-EC-1092 | ERS1704157 | 131 |
| HPRU-EC-1093 | ERS1704158 | 95 |
| HPRU-EC-1094 | ERS1704159 | 62 |
| HPRU-EC-1095 | ERS1704160 | 131 |
| HPRU-EC-1097 | ERS1704161 | 127 |
| HPRU-EC-1098 | ERS1704257 | 131 |
| HPRU-EC-1102 | ERS1704259 | 73 |
| HPRU-EC-1106 | ERS1704261 | 345 |
| HPRU-EC-1107 | ERS1704263 | 95 |
| HPRU-EC-1111 | ERS1704265 | 131 |
| HPRU-EC-1112 | ERS1704266 | 1159 |
| HPRU-EC-1116 | ERS1704267 | 59 |
| HPRU-EC-1122 | ERS1704268 | 404 |
| HPRU-EC-1123 | ERS1704269 | 7518 |
| HPRU-EC-1124 | ERS1704270 | 127 |
| HPRU-EC-1126 | ERS1704271 | 449 |
| HPRU-EC-1127 | ERS1704272 | 69 |
| HPRU-EC-1129 | ERS1704273 | 393 |
| HPRU-EC-1131 | ERS1704274 | 131 |
| HPRU-EC-1132 | ERS1704275 | 62 |
| HPRU-EC-1134 | ERS1704277 | 393 |
| HPRU-EC-1135 | ERS1704278 | 453 |
| HPRU-EC-1136 | ERS1704279 | 131 |
| HPRU-EC-1137 | ERS1704280 | 73 |
| HPRU-EC-1139 | ERS1704281 | 3574 |
| HPRU-EC-1140 | ERS1704282 | 131 |
| HPRU-EC-1141 | ERS1704283 | 131 |
| HPRU-EC-1142 | ERS1704284 | 1193 |
| HPRU-EC-1145 | ERS1704285 | 73 |
| HPRU-EC-1146 | ERS1704286 | 135 |
| HPRU-EC-1148 | ERS1704287 | 131 |
| HPRU-EC-1149 | ERS1704288 | 131 |
| HPRU-EC-1153 | ERS1704290 | 73 |
| HPRU-EC-1154 | ERS1704291 | 62 |
| HPRU-EC-1155 | ERS1704292 | 127 |
| HPRU-EC-1156 | ERS1704293 | 73 |
| HPRU-EC-1161 | ERS1704295 | 131 |
| HPRU-EC-1163 | ERS1704296 | 131 |

| | | |
|---|---|---|
| HPRU-EC-1165 | ERS1704297 | 472 |
| HPRU-EC-1174 | ERS1704298 | 517 |
| HPRU-EC-1175 | ERS1704299 | 7517 |
| HPRU-EC-1177 | ERS1704300 | 131 |
| HPRU-EC-1182 | ERS1704301 | 69 |
| HPRU-EC-1185 | ERS1704302 | 73 |
| HPRU-EC-1186 | ERS1704304 | 131 |
| HPRU-EC-1187 | ERS1704303 | 73 |
| HPRU-EC-1188 | ERS1704305 | 69 |
| HPRU-EC-1189 | ERS1704306 | 131 |
| HPRU-EC-1190 | ERS1704307 | 10 |

87
